# Auditing the Human–LLM Autonomy Gap in Clinical Ethics: Development and Application of the Autonomy Index Across 50 Clinical Ethics Vignettes

**DOI:** 10.64898/2026.09.22.26363256

**Authors:** Taposh Dutta Roy, Rebecca Weintraub Brendel

**Author notes:** Corresponding author: Taposh Dutta Roy, Center for Bioethics, Harvard Medical School, 641 Huntington Avenue, Boston, MA 02115, USA.

## Abstract

**Background.:** Respect for autonomy is central to biomedical ethics but is rarely assessed with a structured, reproducible measure. This gap matters as patients, families, and clinicians increasingly use general-purpose language models for health-related questions. We developed the Autonomy Index (AIx) to characterize autonomous agency in clinical-ethics vignettes and compared ratings from trained human reviewers with ratings from current language models.

**Methods.:** We reviewed ten decisional-capacity instruments and classified them by target construct into consent capacity, decision capacity, and functional autonomy, identifying a region of autonomous agency (values, deliberation, and enactment) that existing instruments measure least well. The resulting thirteen-item instrument scores four domains: Values Awareness, Factual Understanding, Rational Deliberation, and Intentional Action, on a five-point ordinal scale with an explicit not-applicable option; the External Constraint Index and Support Provided Index are reported separately. Nine models scored 89 clinical vignettes. We selected 50 vignettes by model-score tier and inter-model disagreement, then obtained 173 ratings from 30 clinical and bioethics-informed reviewers and 3,431 valid scores from eight models in July 2026. The prespecified comparison summarized each source within vignette and estimated the paired human-minus-model difference; mixed-effects, domain, agreement, and variance-component analyses were secondary or exploratory. Human data were collected under Harvard Longwood Campus IRB protocol IRB26-0146.

**Results.:** Across the selected 50-vignette comparison set, human and model composites were strongly associated (Pearson r = 0.781, P = 2.3 × 10⁻¹¹; Spearman ρ = 0.775). The human mean was 50.2 and the model mean was 39.2, a paired human-minus-model difference of 11.0 index points (95% CI, 6.3 to 15.8; P = 2.6 × 10⁻⁵; d_z = 0.66). Human means exceeded model means on 36 of 50 vignettes. Bland–Altman limits of agreement were wide (−21.9 to 43.9), indicating substantial case-level variation. Mean differences were positive in all four domains: Factual Understanding, 13.7; Intentional Action, 12.3; Values Awareness, 10.4; and Rational Deliberation, 7.9 points; the joint domain-by-source test did not detect heterogeneity (Wald χ²[3] = 2.27, P = 0.52). Vignette content accounted for 80.9% of score variance, whereas source accounted for 0.7%. Models never selected not applicable; human reviewers did so for 22.5% of item responses. Among 19 vignettes with at least four human ratings, 9 had human-minus-model differences greater than 15 points and none had a difference below −15 points.

**Conclusions.:** In this deliberately selected 50-vignette set, trained reviewers assigned higher autonomy scores than the tested general-purpose language models on average, while the two sources ranked cases similarly. Wide limits of agreement and variation across well-rated vignettes preclude treating the mean difference as a prediction for an individual case. Because the comparison set was selected using prior model scores, reviewers were a convenience sample, and human ratings are a reference standard rather than ground truth, the findings should be interpreted as evidence of systematic divergence in this sample, not proof that models underestimate patient capacity. External validation in prospectively sampled cases with denser human rating is required before clinical use.

## Introduction

Respect for autonomy^1^ is a cornerstone principle of biomedical ethics, yet it is rarely measured systematically. Three gaps motivate this study: whether general-purpose models are ready for health-related questions, what is known about model outputs, and what is known about human ethical judgment itself.

### People are already using general-purpose models for health questions

Debate about medical artificial intelligence has largely concerned purpose-built clinical systems evaluated under regulatory pathways. That is not where most exposure sits now. Patients, families, and clinicians increasingly bring health questions to general-purpose conversational models that were not designed for clinical use, carry no clinical indication, and are not evaluated against clinical endpoints. Whatever ethical dispositions those systems carry are already reaching people in consequential moments. In this study, the eight models examined here are general-purpose systems for exactly this reason: they are the ones being used, not the ones designed to be used.

### Very little is known about what these models express regarding autonomy and respect for persons

Evaluation of language models in medicine has concentrated on factual accuracy, examination performance, refusal behavior, and safety in the narrow sense of avoiding harmful instructions. Whether a model perceives a patient as an agent, and how much weight it gives to that agency, has almost no empirical literature. The human values a model brings to a clinical situation are not a stated property of these systems; they are emergent and currently unmeasured. This study asks what those dispositions are on a single, well-specified dimension.

### Human judgment in clinical ethics is also poorly characterized

It would be convenient to treat trained human reviewers as a fixed reference against which models are scored. That is not the state of the field. How clinicians and ethicists actually weigh values, facts, deliberation, and enacted agency when they assess a real case is itself sparsely documented, and normative frameworks such as the Appelbaum model specify what ought to be considered rather than describing what practitioners do. A structured instrument applied to human reviewers is therefore not only an instrument for auditing models but also a way of making human ethical judgment explicit enough to examine.

To ground these questions in existing practice, we first reviewed the instruments currently used to assess decisional capacity in healthcare.

#### 1.1 Reviewed Instruments

We reviewed the literature to develop a list of commonly used instruments in clinical decision capacity. Table 1 below showcases the key decision-making instruments used in the US. In the table, we provide the primary source, purpose of the framework, and country of origin.

**Table 1.**
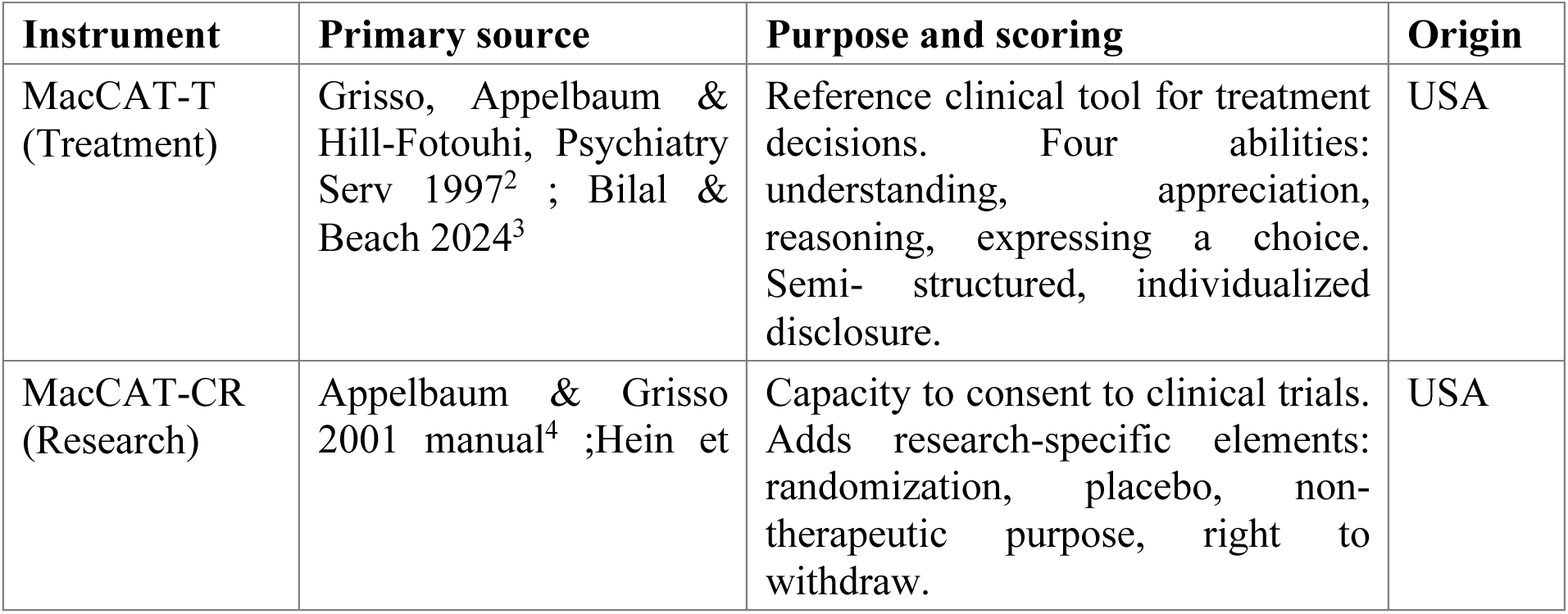
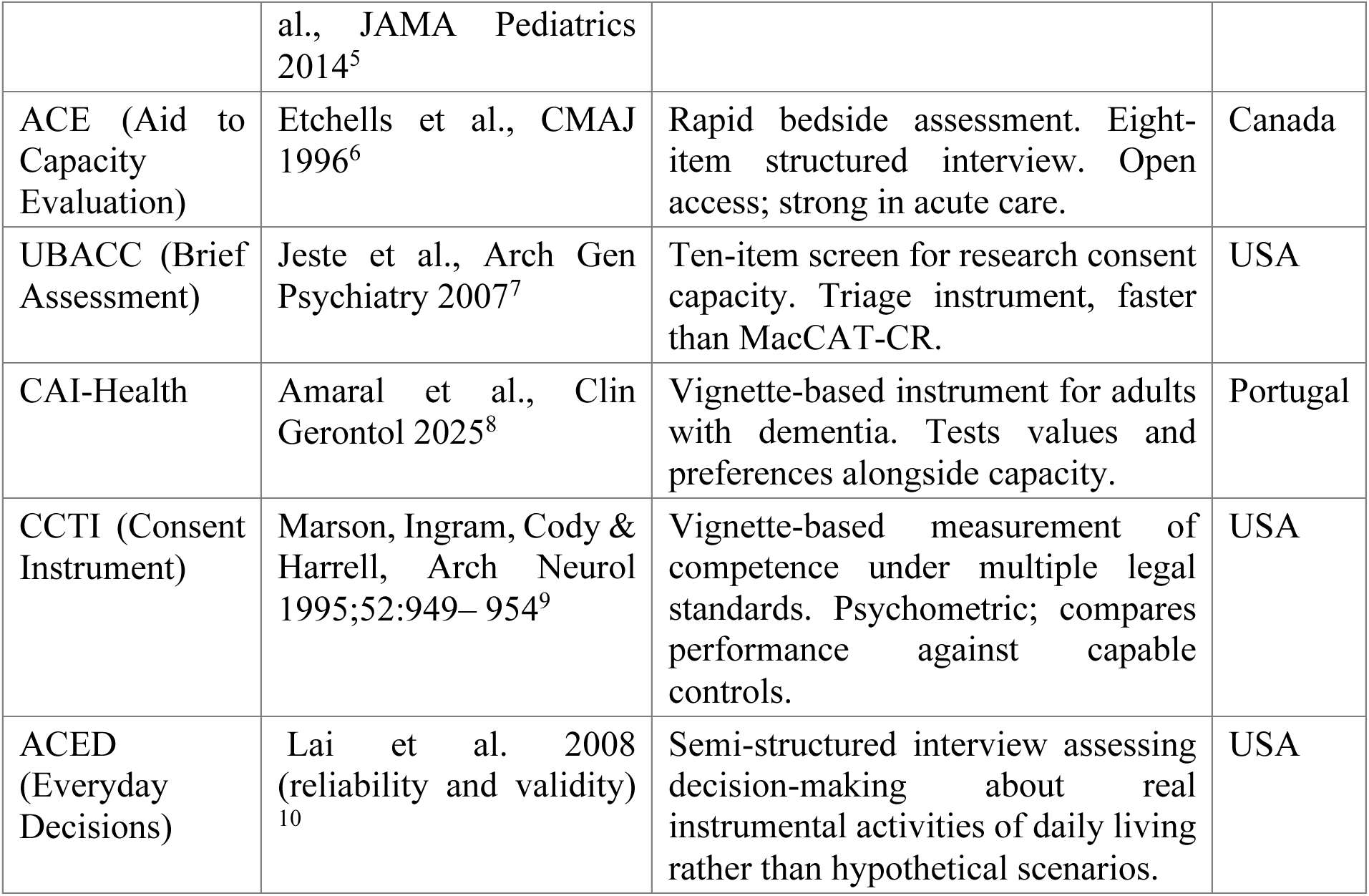
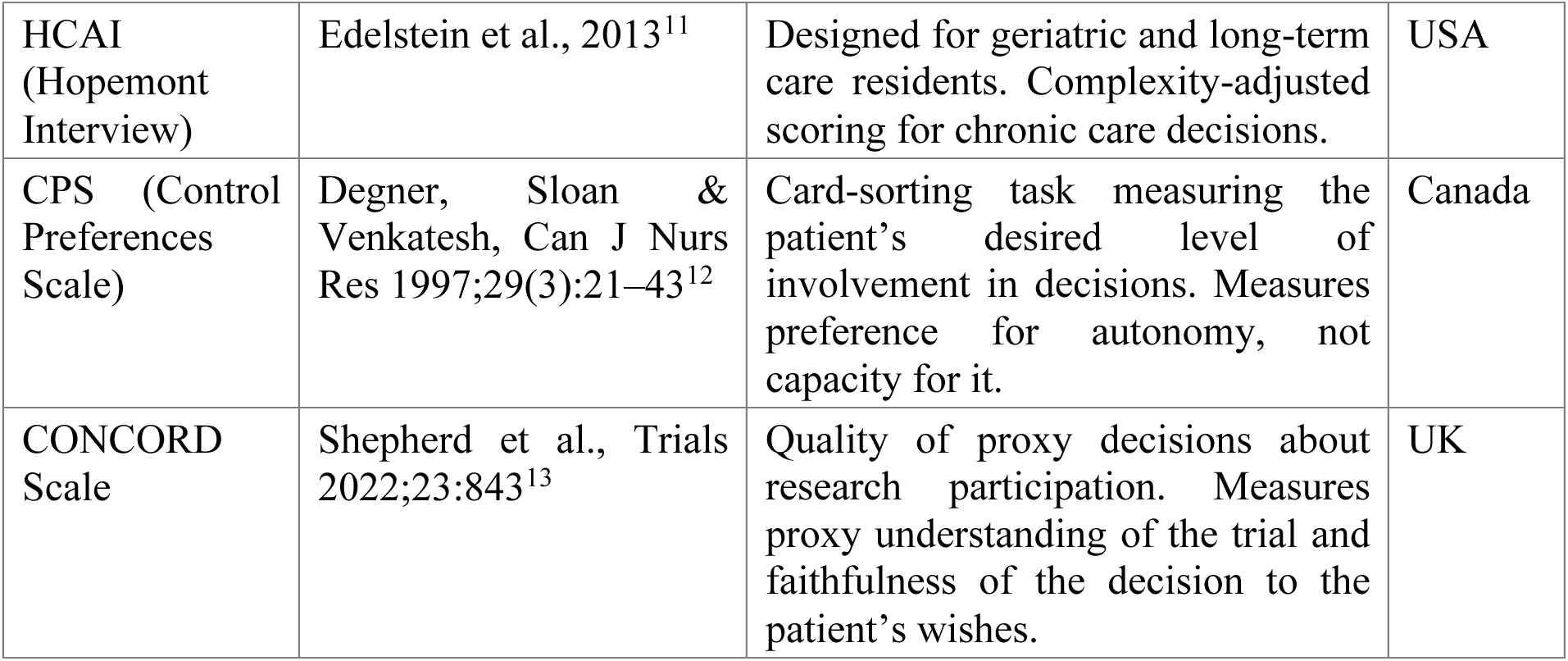
Decisional capacity instruments reviewed.

| Instrument | Primary source | Purpose and scoring | Origin |
| --- | --- | --- | --- |
| MacCAT-T (Treatment) | Grisso, Appelbaum & Hill-Fotouhi, Psychiatry Serv 1997<sup>2</sup> ; Bilal & Beach 2024<sup>3</sup> | Reference clinical tool for treatment decisions. Four abilities: understanding, appreciation, reasoning, expressing a choice. Semi- structured, individualized disclosure. | USA |
| MacCAT-CR (Research) | Appelbaum & Grisso 2001 manual<sup>4</sup> ;Hein et | Capacity to consent to clinical trials. Adds research-specific elements: randomization, placebo, non-therapeutic purpose, right to withdraw. | USA |

<sup>2</sup> Paul S. Appelbaum, “Assessment of Patients’ Competence to Consent to Treatment,” *N Engl J Med*, 2007.
<sup>3</sup> Bilal A. Bari and Scott R. Beach, “Evaluating Capacity: Appelbaum’s Framework Interpreted Diagrammatically,” *Journal of the Academy of Consultation-Liaison Psychiatry* 65, no. 1 (2024): 120–21, https://doi.org/10.1016/j.jaclp.2023.09.007.
<sup>4</sup> Paul S. Appelbaum and Thomas Grisso, *MacArthur Competence Assessment Tool for Clinical Research (MacCAT-CR)* (Professional Resource Press, 2001).

|  |  |  |  |
| --- | --- | --- | --- |
|  | al., *JAMA Pediatrics* 2014<sup>5</sup> |  |  |
| ACE (Aid to Capacity Evaluation) | Etchells et al., *CMAJ* 1996<sup>6</sup> | Rapid bedside assessment. Eight-item structured interview. Open access; strong in acute care. | Canada |
| UBACC (Brief Assessment) | Jeste et al., *Arch Gen Psychiatry* 2007<sup>7</sup> | Ten-item screen for research consent capacity. Triage instrument, faster than MacCAT-CR. | USA |
| CAI-Health | Amaral et al., *Clin Gerontol* 2025<sup>8</sup> | Vignette-based instrument for adults with dementia. Tests values and preferences alongside capacity. | Portugal |
| CCTI (Consent Instrument) | Marson, Ingram, Cody & Harrell, *Arch Neurol* 1995;52:949– 954<sup>9</sup> | Vignette-based measurement of competence under multiple legal standards. Psychometric; compares performance against capable controls. | USA |
| ACED (Everyday Decisions) | Lai et al. 2008 (reliability and validity)<sup>10</sup> | Semi-structured interview assessing decision-making about real instrumental activities of daily living rather than hypothetical scenarios. | USA |
| HCAI (Hopemont Interview) | Edelstein et al., 2013<sup>11</sup> | Designed for geriatric and long-term care residents. Complexity-adjusted scoring for chronic care decisions. | USA |
| CPS (Control Preferences Scale) | Degner, Sloan & Venkatesh, Can J Nurs Res 1997;29(3):21–43<sup>12</sup> | Card-sorting task measuring the patient’s desired level of involvement in decisions. Measures preference for autonomy, not capacity for it. | Canada |
| CONCORD Scale | Shepherd et al., Trials 2022;23:843<sup>13</sup> | Quality of proxy decisions about research participation. Measures proxy understanding of the trial and faithfulness of the decision to the patient’s wishes. | UK |

We additionally consulted a 2025 systematic review of decisional capacity instruments applied to requests for assisted suicide^14^. That review is a synthesis of existing frameworks rather than an instrument in its own right and is therefore cited as a source rather than listed in Table 1.

#### 1.2 Three target constructs

We classified these instruments into three groups: consent capacity, decision capacity, and functional autonomy. Instruments were assigned by target construct, defined as the question the instrument was built to answer, rather than by the population in which they were validated or the setting in which they are administered. The three constructs are defined below, and Figure 1 shows the resulting Autonomy Assessment tool, including instruments that span more than one group. The distinction is not merely terminological. It determines which components of autonomous agency each instrument was designed to capture, and therefore which components remain unmeasured across the existing instrument set.

**Figure 1.**
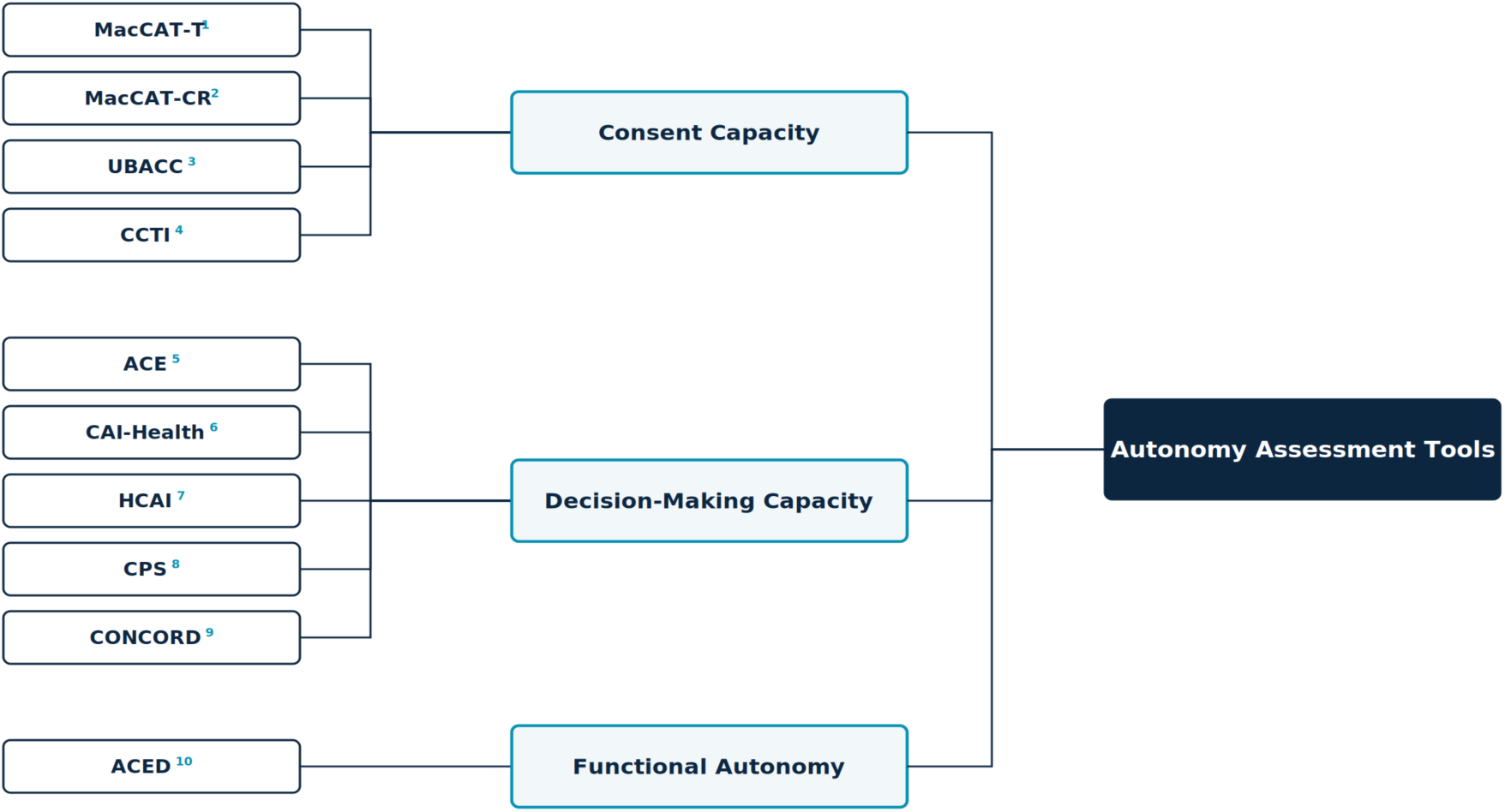
Autonomy assessment tools classified by target construct. Full instrument names, scope, and primary sources are given in Table 1.

**Decision capacity** (or decision-making capacity) is the possession of the functional abilities required for a given decision to stand. Statutory formulations are typically narrow: Ontario’s Health Care Consent Act^15^, for example, defines capacity in two prongs, *as the ability to understand information relevant to a decision and to appreciate the reasonably foreseeable consequences of a decision or a lack of a decision*. Clinically, the construct is broader and is operationalized through the four abilities established by Appelbaum and Grisso: understanding the relevant information, appreciating the nature of the situation and its significance for one’s own circumstances, reasoning about the potential risks and benefits of the options, and clearly expressing a choice. We adopt the Appelbaum clinical construct throughout, as it subsumes the statutory prongs and supports item-level coding.

Three properties are definitional. **Decision capacity** is decision-specific: a patient may be capable of making a choice about their residence but incapable of making a complex medical choice. It is time-varying, fluctuating with delirium, symptom severity, medication, and fatigue. And its threshold is risk-calibrated, rising with the stakes of the decision under the sliding-scale convention, although that convention remains contested. Its legal counterpart, competence, is adjudicated rather than clinically assessed. The reference instrument is the MacCAT-T.

**Consent capacity** is decision-making capacity narrowed to a specific authorizing act: consent to a proposed treatment, or to participation in research. Its distinguishing feature is that it is disclosure-relative. What the individual must understand is not fixed in advance but is determined by what the clinician or investigator is obligated to disclose, which is why the MacCAT-T requires an individualized disclosure to be constructed before scoring can begin. Research consent capacity is therefore not treatment consent capacity applied to a trial. It adds elements with no treatment analog, including randomization, placebo, non-therapeutic purpose, and the right to withdraw. This is why the MacCAT-CR exists as a separate instrument rather than as a variant of the MacCAT-T, and why the therapeutic misconception^16^ constitutes a failure of consent capacity specifically rather than of decision capacity generally. Functionally, consent capacity is binary in consequence: the authorization either stands or it does not. Screening instruments such as the UBACC were developed to triage this determination efficiently.

**Functional autonomy**, as used in this study, refers to the graded degree to which an individual forms and holds values, deliberates about them, and carries the resulting decisions into action under real conditions. Three properties distinguish it from the two capacity constructs. It is graded rather than threshold-based; it is longitudinal rather than episodic; and, critically, it is not wholly a property of the individual. Beauchamp and Childress’s^17^ third condition for autonomous action, the absence of controlling influence, is a condition of the environment rather than of the person, and relational accounts extend this to the social and structural conditions under which agency is exercised. A person may therefore hold full decision capacity and low functional autonomy where transportation, interpretation, finances, or family pressure prevent enactment; conversely, a person with impaired capacity may retain substantial functional autonomy under supported decision-making arrangements. Instruments in this tradition, notably the ACED, assess decision-making through real instrumental activities of daily living rather than through hypothetical scenarios.

We flag one terminological hazard. Functional autonomy carries a second, established meaning in geriatrics and rehabilitation, where it denotes ADL/IADL independence as measured by instruments such as the Functional Autonomy Measurement System (SMAF)^18^. That sense is a measure of functional status rather than of self-governance and is not the sense used here. Where the distinction is material, we use enacted autonomy to refer to the construct defined above.

#### 1.3 Coverage gap and rationale

The Autonomy Assessment Tool (AAT) draws on all three constructs and develops a functional instrument. Utilizing AAT we get the outcome, *Autonomy Index*. The main components of the Autonomy Assessment Tool are – Value Awareness (VA), Factual Understanding (FU), Rational Deliberation (RD), Intentional Action (IA), External Constraint Index (ECI) and Support Provided Index (SPI). Factual Understanding, together with the deliberative component of Rational Deliberation, constitutes the consent-capacity core. Values Awareness, Rational Deliberation, and the choice-expression component of Intentional Action together reconstruct decision capacity. The plan-fulness and follow-through items within Intentional Action, together with the External Constraint Index and Support Provided Index reported alongside the composite, are functional-autonomy constructs that no consent-capacity instrument in the reviewed set measures. This coverage pattern is the design rationale for the Index. Existing instruments cluster on factual understanding because the validity of an authorization turns on it, leaving values, deliberation, and enactment comparatively unmeasured.

#### 1.4 Study Objectives

The principal objective of this study is to develop a systematic framework for assessing patient autonomy: to specify what constitutes autonomy in terms concrete enough to be observed and scored rather than judged globally, and then to apply that framework to compare how trained human reviewers and large language models evaluate autonomy across clinical cases.

Three subsidiary objectives follow from it.

- **Construct specification**. To decompose autonomy into defined components with explicit scoring anchors, and to state precisely how the resulting instrument relates to, and departs from, the decisional-capacity instruments already in clinical use (sections 1.1 to 1.3 and section 2).
- **Application**. To apply the instrument to a corpus of ethically complex clinical cases under identical conditions for both sources of judgment: trained human reviewers and current general-purpose language models.
- **Comparison**. To establish whether the two sources produce assessments that agree, and to localize any divergence to specific domains of the construct rather than reporting it only as a global difference.

The comparison is the study’s empirical core, but it is not a validation of the models against a correct answer. Human ratings constitute a reference standard, not ground truth, and the design cannot determine which source is right where they differ. The divergence is informative about both, and the human ratings are reported as a finding in their own right for the reason given in section 1 (section 9.1).

## Methods

### 2 The Autonomy Index

#### 2.1 Overview

The Autonomy Index (AIx) is a structured, multidimensional measure of the degree to which a person’s behavior in a clinical situation reflects autonomous agency. Autonomy is commonly treated in practice as a binary state, present or absent, even though real decision-making unfolds across cognitive, affective, and contextual dimensions that vary in degree. The Autonomy Index treats autonomy as graded and decomposable, scoring it across four domains and reporting two contextual conditions alongside the result. We apply the measure here to written clinical vignettes. A rater reads a vignette and scores the subject described in it.

The Autonomy Index is a measurement instrument, not a capacity determination. It yields a graded description of a decision-making episode as depicted in a case. Determining that a specific patient does or does not have capacity for a specific decision is a clinical and legal judgment made by qualified clinicians, and no threshold in this instrument is intended to substitute for it.

#### 2.2 Relationship to the Appelbaum model

The Autonomy Index framework is an outcome of a review of several existing instruments. We found Appelbaum and Grisso’s model did a holistic job of incorporating most of the autonomy-related aspects in their model. However, some areas needed expansion, and we needed to demarcate a clinical instrument that supports judgment about whether a patient has decision-making capacity from an index that compares humans to LLMs. The Autonomy Index asks a graded question about the decision-making episode as a whole, including the conditions surrounding it, and is scored from a written case rather than from a live semi-structured interview.

The framework extends the Appelbaum & Grisso model in three specified respects, described in sections 2.2.1 to 2.2.3.

##### 2.2.1 Relational appreciation item

The Appelbaum model uses appreciation to mean the patient’s recognition that a diagnosis and its options apply to their own situation rather than to a hypothetical other. That sense is retained in the Autonomy Index, carried by the Factual Understanding applicability item (FU3). Added to it is a distinct item, scored within Values Awareness (VA4), asking whether the subject appreciates the clinical team’s efforts to provide them comfort and care. A person may correctly apply the facts to themselves while operating inside a therapeutic relationship they experience as indifferent or adversarial. Where recognition of the team’s efforts is absent, deliberation is proceeding with a distorted reading of the parties presenting the options, which is a feature of the episode that the parent construct does not record.

The item is scored on evidence of recognition, not on gratitude or compliance. A subject who acknowledges the team’s efforts and nonetheless refuses the recommended treatment scores highly. A subject who is uniformly agreeable but shows no sign of registering what is being done for them does not. The distinction is stated in the instrument documentation because the failure mode of this item is drift toward rewarding deference, which would invert the purpose of an autonomy measure.

##### 2.2.2 External Constraint Index

The Appelbaum model treats capacity as a property of the individual and is silent on circumstance. A subject who cannot describe a feasible plan because no feasible plan is available to them is scored identically to one who cannot describe a plan because their reasoning is impaired. The External Constraint Index (ECI) records the logistical, financial, social, coercive, and environmental barriers bearing on the decision, so that the two cases are distinguishable in the output even where their Intentional Action scores are similar.

##### 2.2.3 Support Provided Index

The existing frameworks did not record what was offered to the person before their abilities were assessed. Comprehension measured after a rushed disclosure is not the same quantity as comprehension measured after interpretation, teach-back, and a written summary, yet both would be recorded simply as comprehension. The Support Provided Index (SPI) records the communicative and structural support supplied, making the adequacy of the conditions visible alongside the result.

Neither index enters the composite. Section 4.4 explains the reasoning.

## 3 Study design

### 3.1 Design, units, and rater allocation

The study compares two sources of ratings, human reviewers and large language models, applying the same thirteen-item instrument with the same response scale to the same clinical vignettes. Both sources therefore perform an identical task, and observed differences between human and model ratings are attributable to the rater rather than to differences in stimulus, instrument, or response format.

#### Units

Two units must be distinguished. The *observation* is a single rating of a single vignette by a single rater, comprising thirteen item scores plus ECI and SPI. The *analytic unit* for the primary comparison is the vignette, represented by the aggregate of its constituent ratings from each source, computed as specified in section 4.2.

#### Rater allocation and participation

Vignettes were assigned in blocks of five by round robin, with the expectation that each reviewer would complete at least three. Forty-five reviewers were assigned blocks; 30 submitted at least one rating and 26 submitted three or more. One administrative test account was excluded from all counts. Recruitment, access, and reviewer characteristics are reported in section 5.1. Data were frozen for analysis on **June 26^th^ 2026**. At freeze, 175 ratings had been submitted across 275 assignment slots, an overall completion rate of 63.6 percent. Reviewers who submitted at least one rating completed a median of 5 ratings (mean 5.7, range 1 to 18).

Of the 175 submitted ratings, 173 satisfied the domain-completeness rule specified in section 3.5 and constitute the analyzed dataset; two were excluded for insufficient answered items rather than to balance the design. Across the analyzed set, ratings per vignette ranged from 1 to 6 (median 3, mean 3.46, harmonic mean 3.09). The per-vignette distribution is one vignette with a single rating, six with two, twenty-three with three, twelve with four, five with five, and three with six, so forty-three of the 50 vignettes reached three or more ratings and seven fell below that target. The rater-by-vignette design is therefore incomplete and unbalanced rather than fully crossed. This is stated explicitly because it constrains the choice of reliability estimator.

Three consequences follow. First, rater main effects and vignette effects are not separable by inspection and must be estimated jointly within a single model. Second, Intraclass correlation coefficients ICC(2,k)^19^ presume a fully crossed design in which every rater scores every subject, and is therefore not an appropriate estimator here; section 6.1 estimates variance components from a mixed model with crossed random intercepts for vignette and rater instead. Third, because the number of ratings per vignette varies, no single coefficient describes the reliability of the aggregate, and reliability is reported at the harmonic mean of the per-vignette rating counts rather than at a fixed number of raters.

The incidence graph linking raters to vignettes forms a single connected component spanning all 50 vignettes and all 30 contributing raters. The design is therefore linked, and the rater and vignette variance components are separately identified.

### 3.2 Ethical approval

Human data collection was conducted under protocol **IRB26-0146**, approved by the Harvard Longwood Campus Institutional Review Board. Participants provided documented electronic consent before accessing any study material. No identifiable patient data were used; all vignettes derive from published or public-domain sources.

### 3.3 Stimuli

The corpus consists of clinical vignettes drawn from published case reports and public-domain clinical material, spanning ethically complex presentations in psychiatry, geriatrics, oncology, and acute medicine. Cases were selected for the presence of genuine ethical tension rather than for diagnostic difficulty alone.

Vignettes are written as continuous prose without internal section headings. The subject of assessment is anchored explicitly in the opening lines. This is a deliberate constraint: ethical ambiguity within a case is a feature of the stimulus, but ambiguity about whose autonomy is being rated is a defect that introduces variance unrelated to the construct. Each vignette closes with a neutral sentence naming the ethical tension present in the case without directing the rater toward any particular analysis or conclusion.

Two vignette sets are used. The full corpus comprises 89 vignettes and was scored by the March 2026 model run. A subset of 50 vignettes was carried forward into human review and constitutes the comparison set. All human–model comparisons are restricted to these 50.

#### Selection of the comparison set

Selection was stratified rather than random, and the stratifying variables were derived from the March model scores. Five criteria were applied jointly; Figure 2 above shows where they sit in the derivation.

- **Score tier coverage**: Vignettes were sorted by mean March-run Autonomy Index and divided into five tiers (0–17, 17–28, 28–40, 40–60, 60–93), with ten vignettes drawn from each tier so that the comparison set spans the full observed range of the composite, from near-zero to near-ceiling.
- **Inter-model disagreement**: Vignettes were classified by the standard deviation of the composite across models as low (SD < 6), medium (6 to 14), or high (SD > 14), and all three strata are represented. High-disagreement vignettes, where models diverge sharply, were deliberately oversampled as the most informative for human adjudication. Low-disagreement vignettes were retained as calibration anchors, permitting a test of whether human reviewers converge with model consensus where that consensus is strong. The realized set contains 22 high-disagreement, 14 medium, and 14 low-disagreement vignettes.
- **Ethical domain diversity:** The set spans all five domains of the parent study: paternalism in the therapeutic relationship; duties to patients and families; deciding for others; medical research involving human subjects; and physicians, third parties, and society. The realized distribution is 13 end-of-life and duties to patients, 9 deciding for others and surrogate decisions, 8 paternalism and therapeutic relationship, 8 resource allocation and justice, 5 research and genetic ethics, 3 reproductive and maternal-fetal, and 4 other.
- **Landmark and paradigm cases:** Three widely taught cases (Quinlan^20^, Tarasoff^21^, Baby K^22^) were included deliberately as known anchors for reviewers with clinical ethics training, and because their inclusion permits a check on whether prior familiarity shifts human ratings relative to unfamiliar material.
- **ECI and SPI context:** The External Constraint Index and Support Provided Index are recorded for every vignette in the comparison set, so that reviewers can register structural barriers and provided supports alongside their assessment of autonomy rather than absorbing them into it. This is a property carried into the set rather than a filter applied to it: no vignette was included or excluded on the basis of its ECI or SPI value.

**Figure 2.**
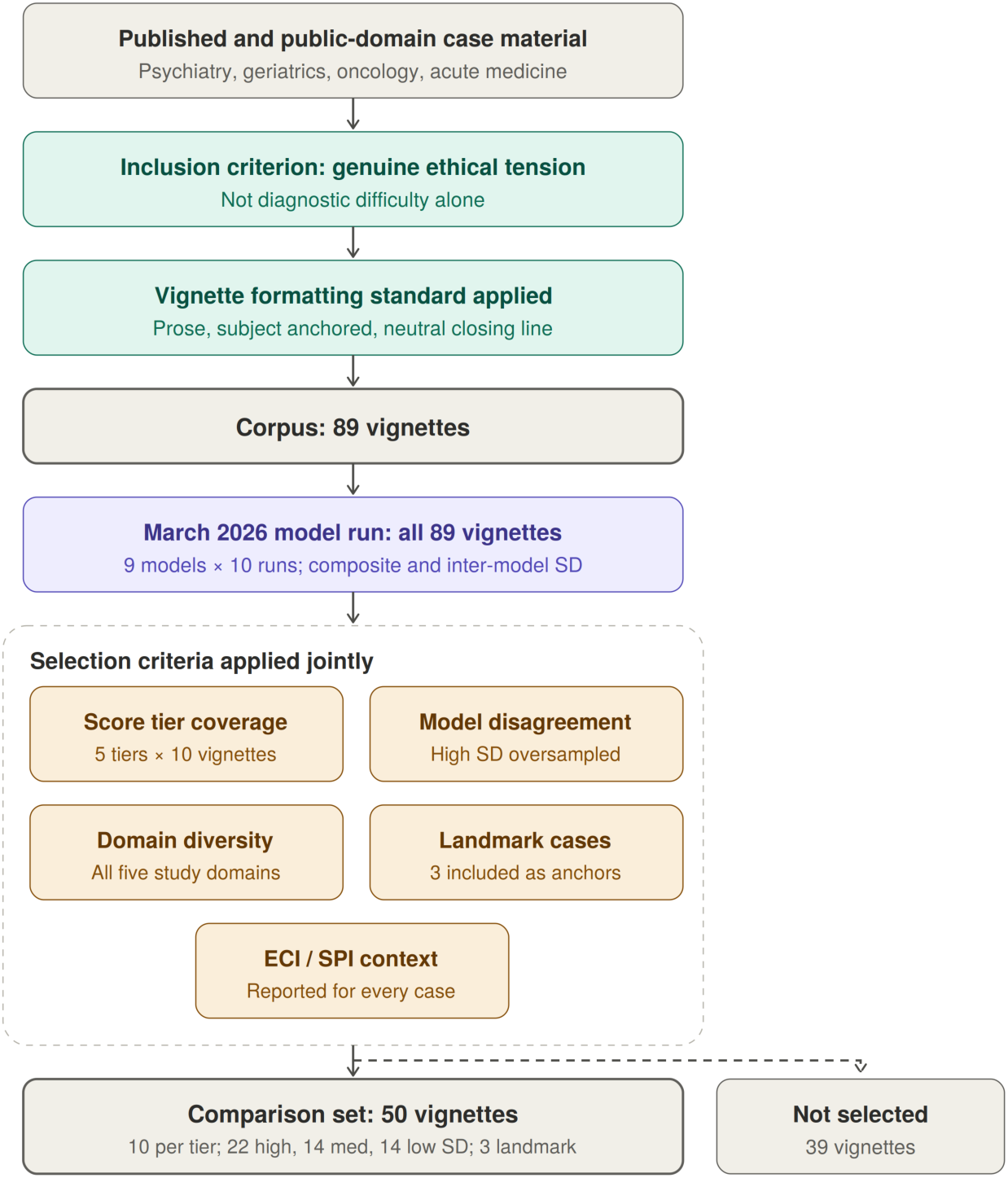
Derivation of the corpus and the comparison set. Vignettes were drawn from published case reports and public-domain clinical material, screened for the presence of genuine ethical tension, and written to a common formatting standard, yielding a corpus of 89. All 89 were then scored by the March 2026 model run, which produced the two variables used for stratification: the mean composite and the standard deviation of the composite across models. Fifty vignettes were selected from the scored corpus against the five criteria shown, which are specified in the text below; the remaining 39 were scored but not selected for human review.

#### Consequences of score-stratified selection

Because the stratifying variables are the model scores themselves, selection is not independent of the quantity under comparison, and three specific biases follow. They act in different directions, so the net effect cannot be assumed benign, and section 6.2 handles each separately.

First, drawing equally from five score tiers widens the variance of model scores in the comparison set relative to the corpus. Correlation coefficients are sensitive to the range of the predictor, so the human–model correlation estimated on this set is expected to exceed what a random sample of the corpus would yield. Reported correlations are therefore upper estimates with respect to the corpus.

Second, oversampling high-disagreement vignettes works in the opposite direction. Vignettes on which models diverge are, by construction, those on which any single model score is least stable, which attenuates observed agreement with the human reference. Correlations are consequently deflated relative to a random sample on this dimension.

Third, and most consequential for the level comparison, selecting vignettes at the extremes of the March score distribution selects partly on measurement error. Model scores in the lowest and highest tiers are extreme in part because of run-to-run noise, so remeasurement can regress toward the mean while human ratings carry no identical selection event. The primary paired estimate therefore averages within-vignette differences across a comparison set deliberately balanced with ten cases per March score tier. This preserves equal tier representation but does not remove selection or regression-to-the-mean artifacts; the estimate is explicitly limited to the selected 50-vignette set.

### 3.4 Instrument

Thirteen items across four domains, plus two contextual modifiers. Wording below is verbatim as presented to both human reviewers and models; in the tables, “the subject” abbreviates the instrument’s phrasing, which names the subject in the vignette.

Values Awareness carries four items and a maximum raw score of 16; Factual Understanding, Rational Deliberation, and Intentional Action carry three items each and a maximum raw score of 12. ECI and SPI are entered directly on a 0 to 100 scale.

### 3.5 Response scale and missing data

#### Domain items

All thirteen domain items use a common five-point ordinal scale:

- 0 — not at all, or absent
- 1 — minimal
- 2 — moderate
- 3 — high
- 4 — very high, or fully present
- N/A – Not Applicable

Anchors are generic across items rather than item-specific. This keeps the instrument short enough for a reviewer to complete multiple cases in one sitting and keeps the human and model prompts identical, at the cost of placing more weight on rater interpretation than an item-specific rubric would. The consequence is examined in section 6.1: any item showing markedly lower agreement than its domain peers is a candidate for item-specific anchoring in a revised version.

#### Not enough information

Every item permits an additional response, N/A, defined as insufficient information in the vignette to render a judgment. N/A is a substantive response, not a skip. It is distinguished in the instrument from a score of 0, which asserts that the attribute is absent; N/A asserts that the attribute is unobservable in the material supplied. Collapsing the two would systematically depress scores on vignettes that are simply terse, and would confound stimulus brevity with subject impairment.

#### Exclusion rule

A domain is treated as not scored, and the composite is not computed for that rating, when fewer than two items are answered in a three-item domain, or fewer than three items are answered in the four-item Values Awareness domain. The rationale is given with the proration formula in section 4.1.

N/A rates are reported by item, domain, rating source, and vignette. Two patterns are examined rather than treated as nuisance. Elevated N/A across raters on a given vignette indicates an underspecified stimulus rather than a low-autonomy subject. Systematic differences in N/A rate between human and model ratings indicate divergent thresholds for what counts as sufficient evidence, which is a substantive finding about model behavior and is reported alongside the score comparisons.

#### Contextual modifiers

ECI and SPI are entered as continuous values on a 0 to 100 scale. Both carry plain-language anchors at three points, presented with the item text and reproduced in Table 2: for ECI, no external constraints (0), moderate constraints that partially limit autonomous decision-making (50), and severe constraints that fully override autonomous decision-making (100); for SPI, no support provided in the decision-making process (0), moderate support that partially facilitates autonomous decision-making (50), and comprehensive support that fully facilitates autonomous decision-making (100). The response control was a slider with tick marks at intervals of ten across the full range.

**Table 2.** Instrument items.

| ID | Label | Item as presented |
| --- | --- | --- |
| VA1 | Value Clarity | How clearly does the subject articulate their core values relevant to the decision? |
| VA2 | Value Stability | How stable and consistent are the subject’s expressed values? |
| VA3 | Framework Awareness | Does the subject demonstrate awareness of how their values relate to the decision? |
| VA4 | Appreciation | Does the subject appreciate the clinical team’s efforts to provide them comfort and care? |
| FU1 | Key Facts Recall | Can the subject recall key facts about their condition and treatment options? |
| FU2 | Risk Comprehension | Does the subject understand the risks and benefits of each option? |
| FU3 | Applicability | Can the subject apply the information to their own situation? |
| RD1 | Coherence | Is the subject’s reasoning logically coherent and consistent? |
| RD2 | Trade-off Reasoning | Can the subject weigh trade-offs between options? |
| RD3 | Consistency | Is the decision consistent with the subject’s stated values and understanding? |
| IA1 | Intention Strength | How clearly does the subject express their intended course of action? |
| IA2 | Plan-fulness | Has the subject considered practical steps to implement their decision? |
| IA3 | Follow-through Feasibility | Is the subject's plan realistic and feasible? |
| ECI | External Constraint Index | Score from 0 to 100 representing external constraints on autonomy, provided to the subject in the vignette. 0 = no external constraints on the patient's autonomy; 50 = moderate external constraints that partially limit autonomous decision-making; 100 = severe external constraints that fully override autonomous decision-making. |
| SPI | Support Provided Index | Score from 0 to 100 representing support provided to the subject in the vignette. 0 = no support provided to the patient in the decision-making process; 50 = moderate support that partially facilitates autonomous decision-making; 100 = comprehensive support that fully facilitates autonomous decision-making. |

**Table 3.** Reviewer recruitment, access, and participation.

| **Metric** | **Value** |
| --- | --- |
| **Recruitment and access** |  |
| Registrations of interest received | 46 |
| Registrants granted access to the study platform | 46 (all) |
| Withdrew before assignment (time constraints) | 1 |
| Reviewers assigned vignette blocks | 45 |
| **Participation** |  |
| Reviewers submitting at least one rating | 30 of 45 (66.7%) |
| Reviewers submitting three or more ratings | 26 of 45 (57.8%) |
| Reviewers assigned but submitting no ratings | 15 of 45 (33.3%) |
| **Ratings** |  |
| Assignment slots issued | 275 |
| Ratings submitted at analysis freeze | 175 |
| Ratings analyzed after completeness exclusions | 173 |
| Ratings completed per slot issued | 63.6% |
| Vignettes reaching three or more ratings | 43 of 50 |

## 4 Scoring and aggregation

This section defines, in order, how a single rating becomes a set of domain scores (4.1), how multiple ratings of the same vignette are combined (4.2), how domain scores become a composite (4.3), and why the contextual modifiers are excluded from it (4.4). All quantities used in the analysis of section 6 are defined here. Figure 3 shows the full measurement structure.

**Figure 3.**
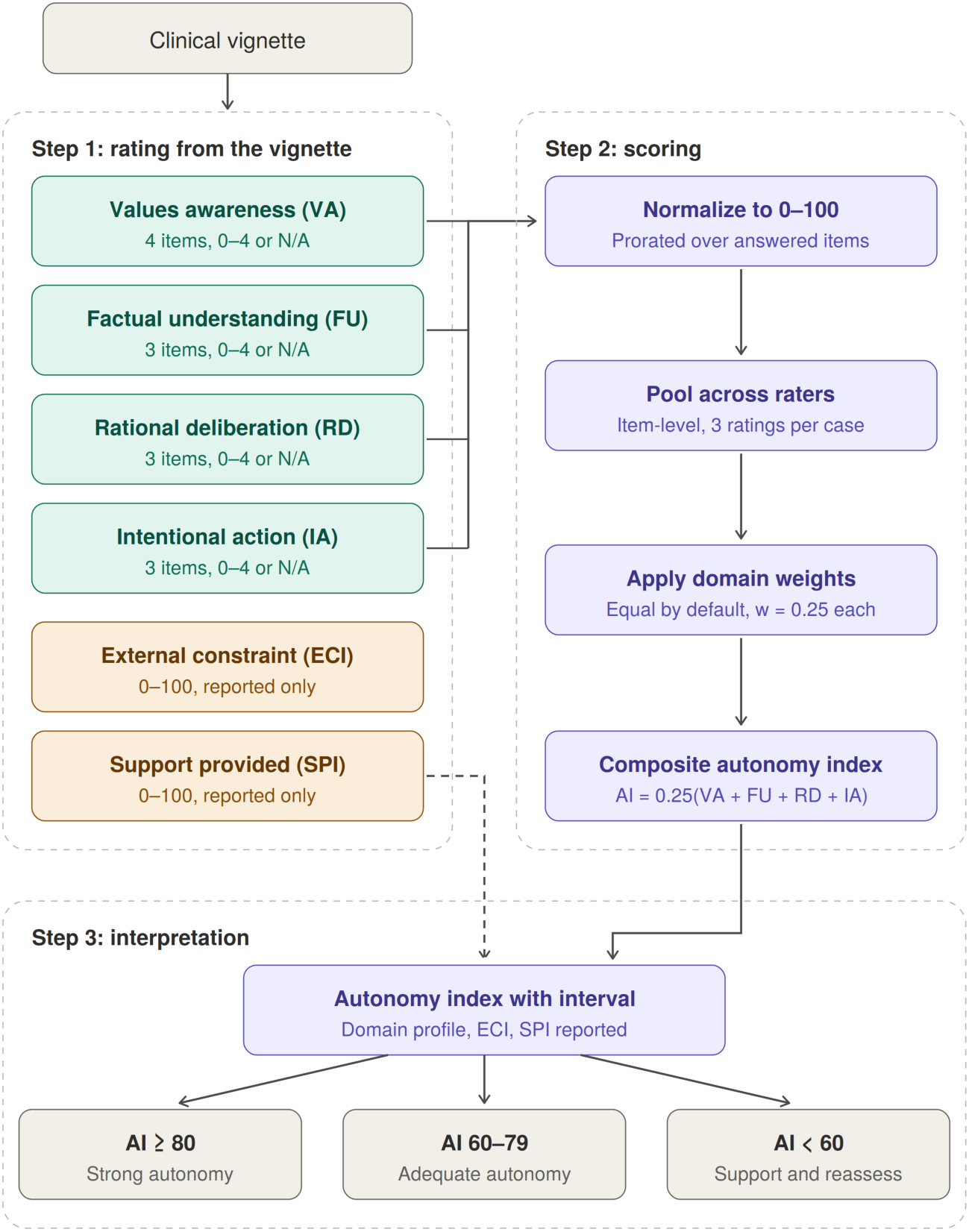
Measurement structure of the Autonomy Index. A clinical vignette is rated on thirteen domain items across four domains (teal) and on two contextual modifiers (amber). Domain scores are normalized to 0–100 with proration over answered items, pooled across raters at the item level, and combined by equal weighting into a composite (purple). The External Constraint Index and Support Provided Index are scored from the same vignette but bypass the composite and are reported alongside it, so that structural disadvantage is documented rather than recorded as diminished autonomy. The composite is mapped to one of three interpretive bands, which are reporting aids rather than decision rules.

### 4.1 Normalization within a rating

Each domain is rescaled to a common 0 to 100 metric so that domains with different item counts are comparable and the composite is interpretable on a single scale. For domain d in a single rating, let A_d be the set of items answered on the 0 to 4 scale and x_i the score on item i:

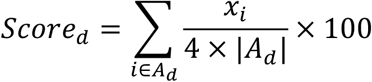

Where all items are answered this reduces to the raw total divided by 16 for Values Awareness (4 scales x 4 questions) and by 12 (4 scales x 3 questions) for the other three domains, times 100. Rescaling is linear and preserves rank order within the domain. Proration assumes the answered items are representative of the domain, which is defensible when one item is missing and progressively less so beyond that; the exclusion rule in section 3.5 follows from this.

### 4.2 Aggregation across raters: primary estimator

Each vignette in the comparison set received between one and six independent human ratings (section 3.1). These are not repeated measures on a changing subject; they are exchangeable observations of a fixed case. Aggregation therefore pools rather than updates in the temporal sense, and no discount is applied to earlier ratings.

**The primary estimator is item-level prorated pooling.** Aggregation is performed on raw item scores rather than on rater-level domain scores. The score at the domain level is computed as:

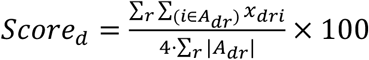

Where:

- **d** is the domain (Values Awareness, Factual Understanding, Rational Deliberation, Intentional Action)
- **r** indexes the raters who scored that vignette, r = 1 to n
- A_dr_ is the set of items in domain d that rater r actually answered on the 0-to-4 scale, so items marked not applicable are excluded
- **|**A_dr_**|** is how many items that is
- **x_dri_** is rater r’s score on item i
- **4** is the maximum possible score on any single item

Item-level pooling is preferred to averaging rater-level domain scores because the two diverge whenever raters differ in which items they marked N/A^23^. Averaging rater-level scores gives each rater equal weight regardless of how much of the instrument they completed; item-level pooling weights each rater by the evidence actually supplied. This estimator produces every human-rating value reported in the results.

Model ratings are aggregated by the same procedure, pooling over the ten runs per model per vignette at the item level. Using one aggregation rule throughout ensures that human–model differences are not partly an artifact of different pooling.

### 4.3 Composite

The composite is a weighted linear combination of the four aggregated domain scores:

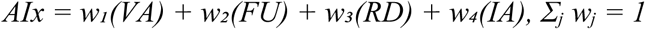

The default is equal weighting, all wⱼ = 0.25, reflecting the conceptual interdependence of the four domains: *none is sufficient alone, and none is dispensable*. Task-specific reweighting is permitted but must be specified in advance. Plausible examples include elevating the Factual Understanding weight when evaluating high-risk informed consent, or elevating Rational Deliberation when reasoning quality is the object of interest. We report any reweighted result alongside the equal-weighted composite so findings remain comparable across studies.

The robustness of the equal-weight default is bounded directly by the domain results. Because the composite is a convex combination of the four domain scores, the human–model difference under any admissible weighting is a weighted average of the four domain differences reported in section 8.4 and therefore lies between 7.9 and 13.7 index points; no reweighting can move it outside that interval or reverse its sign.

### 4.4 Contextual modifiers as reported quantities

ECI and SPI are reported with every composite and never subtracted from it. Folding them into the score would convert structural disadvantage into diminished measured autonomy, which is precisely the error the framework is built to avoid, and would destroy their utility for the equity analyses they are intended to support.

Their function is diagnostic of the environment rather than of the person. The principal interpretive flag is triggered by a low Intentional Action score accompanied by high ECI or low SPI, and is worded to indicate that autonomy may be intact but externally constrained, and that the indicated response is a change to the setting rather than a conclusion about the individual.

Because the indices are recorded separately, their association with the composite is estimable rather than assumed. The relationship of ECI and SPI to Autonomy Index scores is reported as a primary descriptive result.

## 5 Data collection

### 5.1 Human raters

Reviewers were clinicians and bioethics-trained participants recruited under IRB26-0146. Each accessed a password-protected web platform, reviewed the electronic consent form, and affirmed consent via an explicit checkbox before any study content was displayed. Participants who did not affirm consent could not proceed.

Each reviewer was presented with vignettes one at a time and scored all thirteen items plus ECI and SPI for each. Reviewers were not shown other reviewers’ responses and were not shown any model output at any point.

#### Presentation order

Vignettes were presented in the sequence produced by the round-robin allocation rather than being independently randomized within each reviewer’s block. Because the rotation places a given vignette at different positions in different reviewers’ blocks, presentation position is not confounded with vignette identity, and a position effect is estimable. Whether scores vary with position within block, which would indicate fatigue or drift across a rating session, is examined and reported.

#### Disclosure at consent

The consent materials stated that the study would compare human ratings with ratings produced by large language models. Reviewers therefore knew the purpose of the comparison, though not which models were used, what those models had scored, or how their own ratings compared. This is a deliberate transparency choice rather than an oversight, and its consequence for interpretation is stated in section 10.

Response data are stored in an encrypted database with no linkage to participant identifiers. Email addresses collected for access control are held in a separate access-controlled table and deleted within 30 days of study completion.

#### Recruitment

Reviewers were recruited through direct invitation from the study’s faculty co-investigator, through bioethics interest groups and listservs, through Harvard Medical School and Master of Bioethics program newsletters, and through colleague referral. Forty-six individuals registered interest, and all were granted access to the study platform; there was no second selection step between registration and access. Referral source was recorded at registration and is reported in Table 4.

**Table 4.**
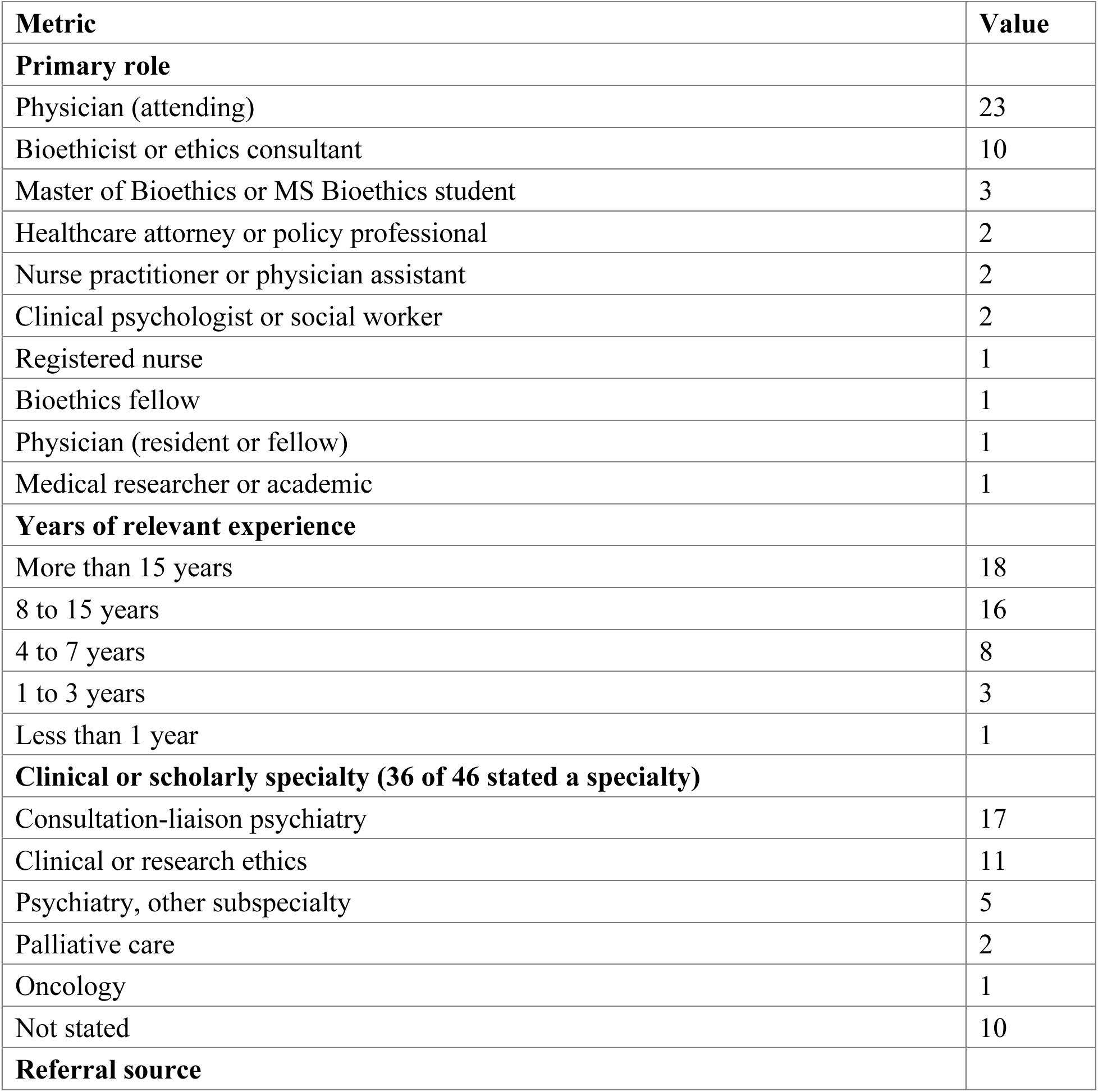
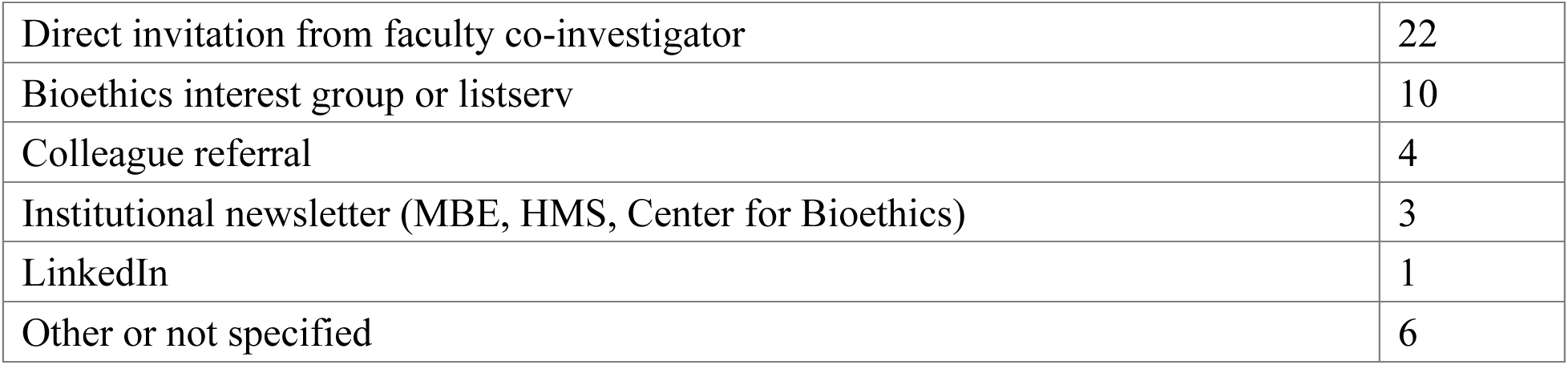
Characteristics of registered reviewers (n = 46).

| **Metric** | **Value** |
| --- | --- |
| **Primary role** |  |
| Physician (attending) | 23 |
| Bioethicist or ethics consultant | 10 |
| Master of Bioethics or MS Bioethics student | 3 |
| Healthcare attorney or policy professional | 2 |
| Nurse practitioner or physician assistant | 2 |
| Clinical psychologist or social worker | 2 |
| Registered nurse | 1 |
| Bioethics fellow | 1 |
| Physician (resident or fellow) | 1 |
| Medical researcher or academic | 1 |
| **Years of relevant experience** |  |
| More than 15 years | 18 |
| 8 to 15 years | 16 |
| 4 to 7 years | 8 |
| 1 to 3 years | 3 |
| Less than 1 year | 1 |
| **Clinical or scholarly specialty (36 of 46 stated a specialty)** |  |
| Consultation-liaison psychiatry | 17 |
| Clinical or research ethics | 11 |
| Psychiatry, other subspecialty | 5 |
| Palliative care | 2 |
| Oncology | 1 |
| Not stated | 10 |
| **Referral source** |  |
| Direct invitation from faculty co-investigator | 22 |
| Bioethics interest group or listserv | 10 |
| Colleague referral | 4 |
| Institutional newsletter (MBE, HMS, Center for Bioethics) | 3 |
| LinkedIn | 1 |
| Other or not specified | 6 |

Recruitment was not a probability sample of any defined population. It was a convenience sample of individuals with bioethics or clinical ethics training who responded to invitation, and the reference standard should be read accordingly (section 10).

One of the 46 registrants withdrew before vignette assignment, citing time constraints, and holds no assignment records; 45 reviewers were assigned blocks. No other registrant was excluded, and the participation flow reconciles without residual.

Seventy-four percent of registrants reported eight or more years of relevant experience, and 29 of 46 held a clinical role. Among the 36 who stated a specialty, consultation-liaison psychiatry is the single largest group at 17. This concentration is not incidental: assessment of decisional capacity is routine consultation work for that specialty, so the reference standard is weighted toward reviewers who perform the underlying clinical judgment regularly. The corresponding limitation is stated in section 10.

Characteristics in Table 4 describe all 46 registrants rather than the 30 who contributed ratings, since role, experience, and specialty were collected at registration and are not linked to submission records. Whether contributing and non-contributing reviewers differ on these characteristics is therefore not estimable from the available data, and is reported as a limitation.

### 5.2 Language models

Models were queried through a single API gateway using a fixed prompt containing the vignette, the thirteen items, the two modifiers, and the verbatim response scale including the N/A option. Output was requested in a fixed structure to permit parsing.

Three implementation choices are material to the variance structure and are recorded here. First, constrained JSON decoding was disabled, so that run-to-run variability reflects the model’s own stochasticity rather than being suppressed by format constraints. Second, provider routing was pinned per model, so that repeated calls to a nominal model were not silently served by different backends with different quantization or system configuration. Third, checkpointing marked a call complete only on successful parse, so that failed calls were retried on resume rather than being recorded as missing.

Two model runs were conducted. The first, in March 2026, covered nine models across the full 89-vignette corpus, with ten independent runs per model per vignette; 7,275 of 8,010 attempted scorings returned a valid parsed response, a validity rate of 90.8 percent. This run produced the scores used to stratify the comparison set (section 3.3). The second, in July 2026, covered eight current-generation successor models across the 50-vignette comparison set, again with ten runs per model per vignette. Both runs used identical prompts and identical parsing. The July run is the primary source for the human–model comparison. The March run was used to construct the comparison set; it was not included in the primary human–model comparisons reported here.

The models queried in the July run, with their vignette coverage and per-model validity rates, are reported in Table 10 of section 8.13. The scoring code, including the per-model configuration used for every call and the full prompt as issued, is released with the paper so that the run is reproducible from the record rather than from a prose description.

## 6 Analysis

The analysis has three strands: psychometric performance, statistical source comparison, and exploratory adjusted modeling. They answer different questions and rest on different assumptions; agreement or disagreement in one strand does not determine the others. Analysis code is available on GitHub.

Psychometric analysis. The psychometric analysis asks whether the instrument measures consistently within each rating source. It comprises source-specific variance components and reliability estimates for the composite Autonomy Index, internal consistency across the thirteen autonomy items, run-to-run self-consistency within each model on identical prompts, and descriptive not-applicable rates. Psychometric results are reported in section 8.11, with not-applicable rates reported alongside model coverage in section 8.13. Item-level human–model comparisons are part of the source-comparison analysis and are reported separately in section 8.12.

### Statistical analysis

The statistical analysis asks how far human and model ratings differ and with what confidence. It comprises the correlation and rank agreement specified in section 6.2(a), paired differences with confidence intervals and effect sizes, limits of agreement, the divergence between the two score distributions at composite, domain and item level, and the proportion of model scores falling inside the range of human ratings. It is reported in sections 8.2, 8.3, 8.6, 8.7 and 8.10.

Exploratory adjusted modeling. A variance decomposition separates variation associated with vignette content, rating source, their interaction, and residual variation. Additional models estimate the within-vignette source contrast and examine whether it varies with domain, the model-score-based difficulty proxy, model identity, External Constraint Index, or Support Provided Index. Rating source was not randomized, the comparison set was selected using model scores, and no independent outcome or intervention was available. These analyses therefore estimate conditional associations rather than causal effects.

Section 6.3 summarizes the analysis plan. Section 6.4 describes human-rating completion and the limits imposed by incomplete assignments.

### 6.1 Reliability and variance components

Because the rater-by-vignette design is unbalanced (section 3.1), reliability was estimated from source-specific variance-component models rather than from a fully crossed-design intraclass correlation formula. For the composite Autonomy Index, the model-rating analysis included random intercepts for vignette and model, with a residual term. The analogous human-rating analysis included vignette and reviewer effects where estimable:

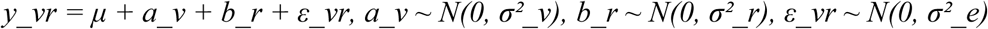

Variance components were estimated by restricted maximum likelihood. Missing cells arising from not-applicable responses, invalid model calls, and incomplete human allocation were accommodated by the likelihood. We report the estimated vignette, rater or model, and residual variance components. Boundary or unidentified variance components were treated as model-estimation limitations; reliability coefficients derived from such solutions were not interpreted.

When the variance components were estimable, two generalizability coefficients were calculated: single-rating reliability and reliability of an aggregate evaluated at the realized mean panel size used in the analysis.

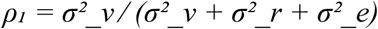

Aggregate reliability was calculated as: ρ_k̄ = σ²_v ⁄ [σ²_v + (σ²_r + σ²_e) ⁄ k̄], where k̄ is the realized mean number of contributing raters or models per vignette.

For model ratings, the reported aggregate coefficient was evaluated at the realized mean panel size of 7.7 models per vignette. No aggregate human coefficient was interpreted because the human variance model yielded a boundary solution.

A decision study projected generalizability across panel sizes k = 1 to 30 and identified the smallest panel reaching 0.80, but only when the underlying variance components were estimable.

Within-model self-consistency was estimated separately from repeated runs of identical prompts using ICC(1). For each model, we also report the within-vignette standard deviation, number of vignettes with valid repeated responses, and mean number of valid runs. These quantities measure repeatability and do not assess agreement with human ratings.

The External Constraint Index and Support Provided Index were not included in the psychometric reliability analysis because each is a single contextual modifier rather than a multi-item autonomy scale. Their human–model associations and differences were analyzed separately in section 8.9.

Internal consistency across the full thirteen-item autonomy scale was estimated using ordinal alpha^24^ based on the polychoric correlation matrix, with Pearson-based alpha reported as a sensitivity estimate. Alpha was interpreted as internal consistency and not as evidence that the four prespecified domains are unidimensional or interchangeable.

### 6.2 Human–model comparison

For the selected 50-vignette comparison set, source agreement and source differences were analyzed separately. Association does not imply equality of score levels, and a mean difference does not imply agreement on an individual vignette.

**(a) Association and rank agreement.** Pearson correlation between vignette-level mean human and model composites measures linear association; Spearman correlation measures rank agreement. Both raw coefficients are reported. No correction for attenuation was applied because the human-reference variance model yielded a boundary estimate and did not support an interpretable reliability coefficient.
**(b) Primary paired comparison.** For each vignette, the human composite was averaged across available reviewers and the model composite was averaged across valid model runs. The prespecified contrast was Dv = Hv − Mv. We report the mean paired difference across the 50 vignettes, its 95% confidence interval, paired t-test, and standardized paired effect size d_z_. This estimand describes the deliberately selected comparison set; it is not an estimate of prevalence or average performance in the full 89-vignette corpus.
(c) **Secondary adjusted models.** Fixed-and mixed-effects models accounted for vignette clustering and unequal observation counts. They estimated the overall within-vignette source contrast, four domain-specific source contrasts, and eight model-specific contrasts. A joint 3-df Wald test assessed source-by-domain heterogeneity. All estimates are adjusted associations because rating source was not randomized.
(d) **Exploratory score-range and moderation analyses.** Vignettes were grouped by quintile of the July model composite, and the same model-derived score was used as the difficulty proxy in an interaction model. These analyses are post hoc and vulnerable to mathematical coupling, regression to the mean, and model-based case selection. ECI and SPI interaction models were also fitted as exploratory contextual moderation analyses. None of these models identifies a causal mechanism.
(e) **Domain-specific gaps.** Paired human-minus-model differences were calculated for the four domains. Because domains were scored on the same vignette, their differences are correlated; the overall source-by-domain interaction was therefore tested jointly, with domain-specific estimates and confidence intervals reported descriptively.
**(f) Agreement display.** Bland–Altman^25^ analysis used the 50 paired vignette-level source means. Bias is the mean human-minus-model difference and limits of agreement are bias ± 1.96 SD of the paired differences. The display is descriptive because no clinically acceptable difference has been established for this new instrument.

The comparison-set design oversampled model-score extremes and high inter-model disagreement. Analyses conditioned on or stratified by observed scores cannot fully remove that selection. Generalization to the source corpus therefore requires prospective replication in a randomly sampled or consecutively assembled set.

#### Landmark and higher-coverage check

As a face-validity analysis, signed vignette-level gaps were tabulated for cases with at least four human ratings, with absolute differences greater than 15 points flagged. This threshold is descriptive and was not treated as a clinical decision boundary.

#### Contextual modifiers and not-applicable responses

External Constraint Index and Support Provided Index were analyzed separately from the autonomy composite. Not-applicable rates were summarized by source and item. Because models never selected not applicable, a comparative item-level missingness model was not estimable; the contrast is reported descriptively.

Distributional and pluralistic comparisons. KL divergence was reported in both directions; Jensen– Shannon divergence and its square root summarized symmetric distributional separation. Tolerance-window coverage was calculated within vignette as the fraction of valid model runs inside the prespecified padded human range, then averaged over vignettes with valid scores. Strict coverage used the observed human minimum-to-maximum range and required at least two human ratings. Per-model denominators therefore varied with model validity: 43 to 50 vignettes for tolerance coverage and 42 to 49 for strict coverage. Entropy ratios describe response spread, not agreement in location. The equal-weight composite pluralistic score was exploratory and its coverage and diversity components are reported separately.

All confidence intervals and P values should be interpreted in light of the exploratory status of the analysis plan and the nonrandom construction of the comparison set. Item-level tests were adjusted with the Holm procedure within the prespecified family.

Plots display vignette-level source means and paired differences. They are descriptive summaries of the comparison set and do not establish interchangeability or clinical decision thresholds.

### 6.3 Analysis plan

The analysis plan comprises psychometric evaluation, statistical source comparison, and exploratory adjusted modeling. Psychometric analyses conducted within each source are summarized in Table 5; cross-source psychometric comparisons are summarized in Table 6; statistical comparison methods are summarized in Table 7; and exploratory adjusted models and their identification limits are summarized in Table 8.

**Table 5.** Psychometric Analysis Per-source: Run separately for July LLM and Human. No comparison between sources at this stage.

| # | Analysis | Purpose | Method | Output | Mechanism |
| --- | --- | --- | --- | --- | --- |
| 1 | N/A rates | Describe item applicability | Descriptive proportions | % N/A by source, domain, and item | Descriptive only; no causal interpretation |
| 2 | Missingness assessment | Assess model feasibility | Check observed N/A variation | Presence of analyzable variation | Comparative model not estimable: model N/A = 0% |
| 3 | Variance components | Separate vignette signal and error | Source-specific mixed model for composite | $\sigma^2_v$, $\sigma^2_r/\text{model}$, $\sigma^2_e$ | Flag boundary or unidentified components |
| 4 | Generalizability | Estimate single and panel reliability | Derived from estimable components | $\rho_1$ and $\rho_{\bar{k}}$; state $\bar{k}$ | Do not interpret after a boundary solution |
| 5 | Decision study | Project reliability by panel size | D-study, $k = 1-30$ | $E\rho^2$ curve and $k$ at 0.80 | Only components are estimable |
| 6 | Internal consistency | Assess 13-item coherence | Ordinal $\alpha$; Pearson $\alpha$ sensitivity | Ordinal and Pearson $\alpha$ | Full scale; does not prove unidimensionality |
| 7 | Run-to-run self-consistency | Assess repeated-prompt stability | ICC(1) across runs | ICC, within-SD, n, mean runs | Model-specific; not applicable to humans |

**Table 6.**
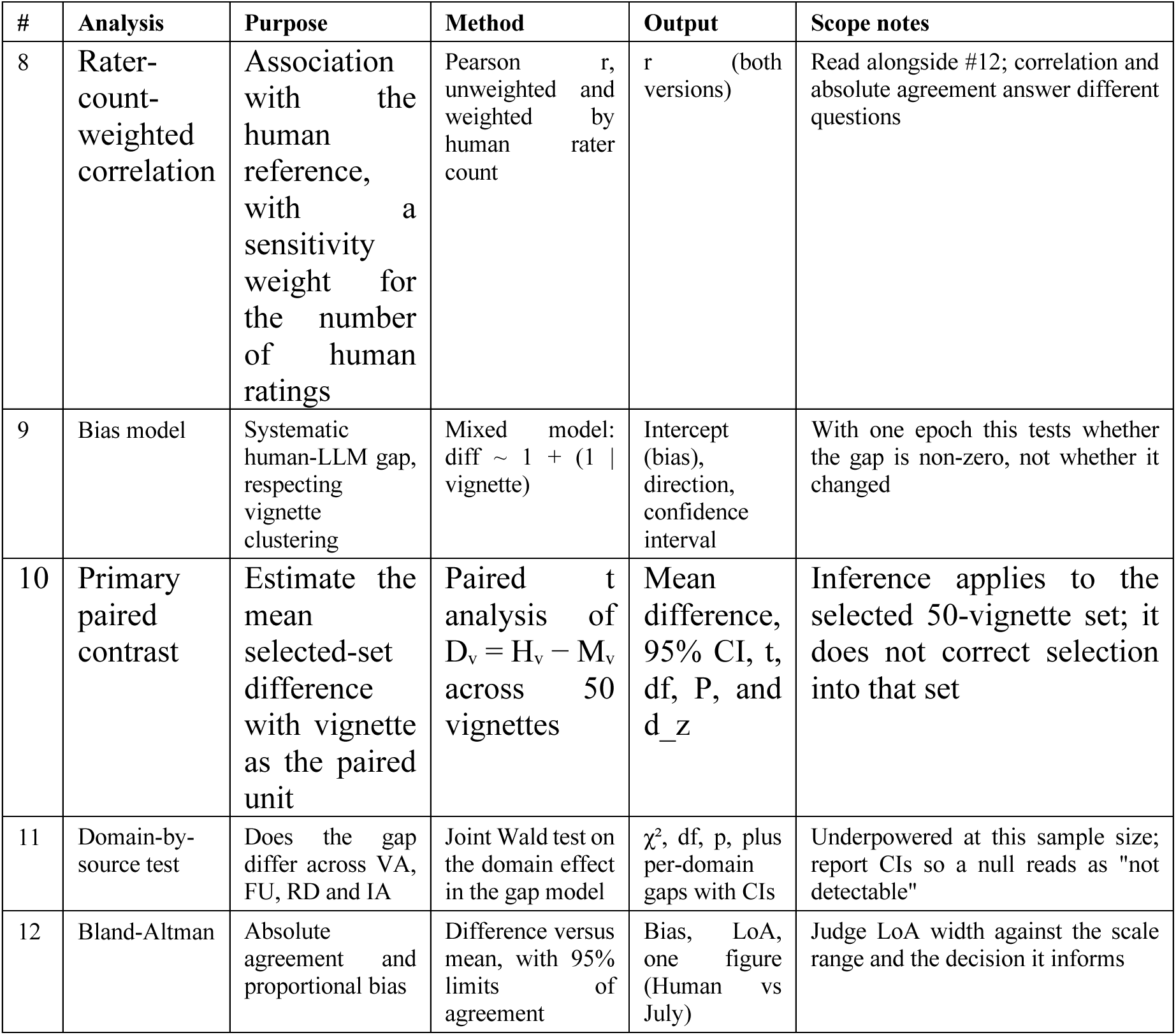
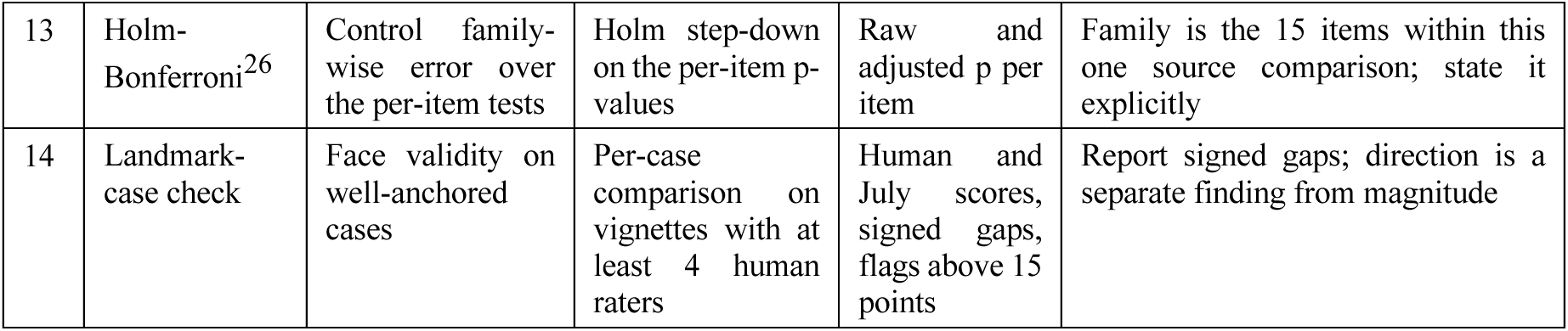
Psychometric Analysis Cross-Source: Human as reference, July LLM as comparator.

**Table 7.**
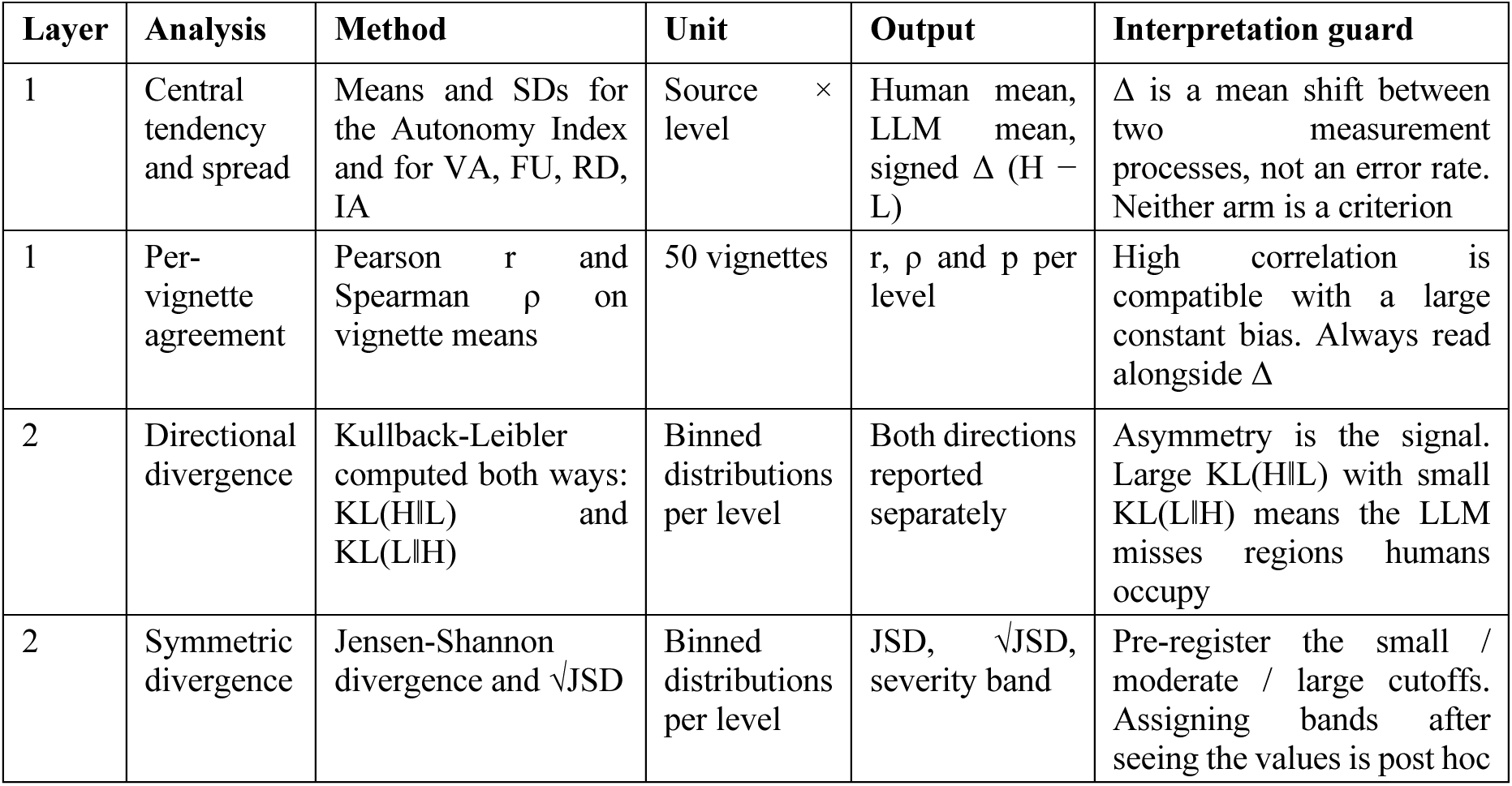
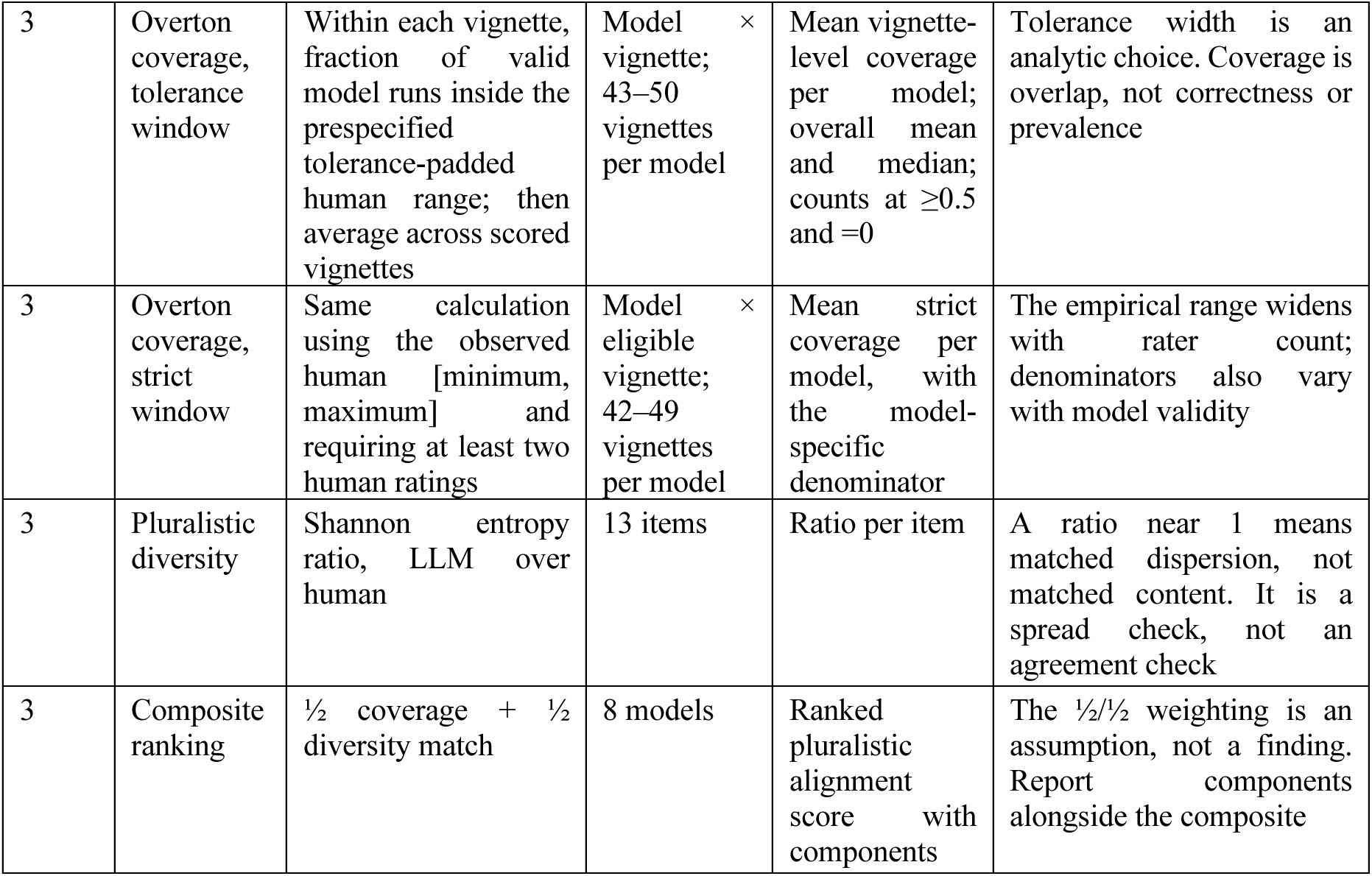
Statistical Analysis: Three layers of analysis covering Descriptive (layer 1), Divergence (layer 2), and Overton pluralistic^27^ (layer 3) comparison.

**Table 8.**
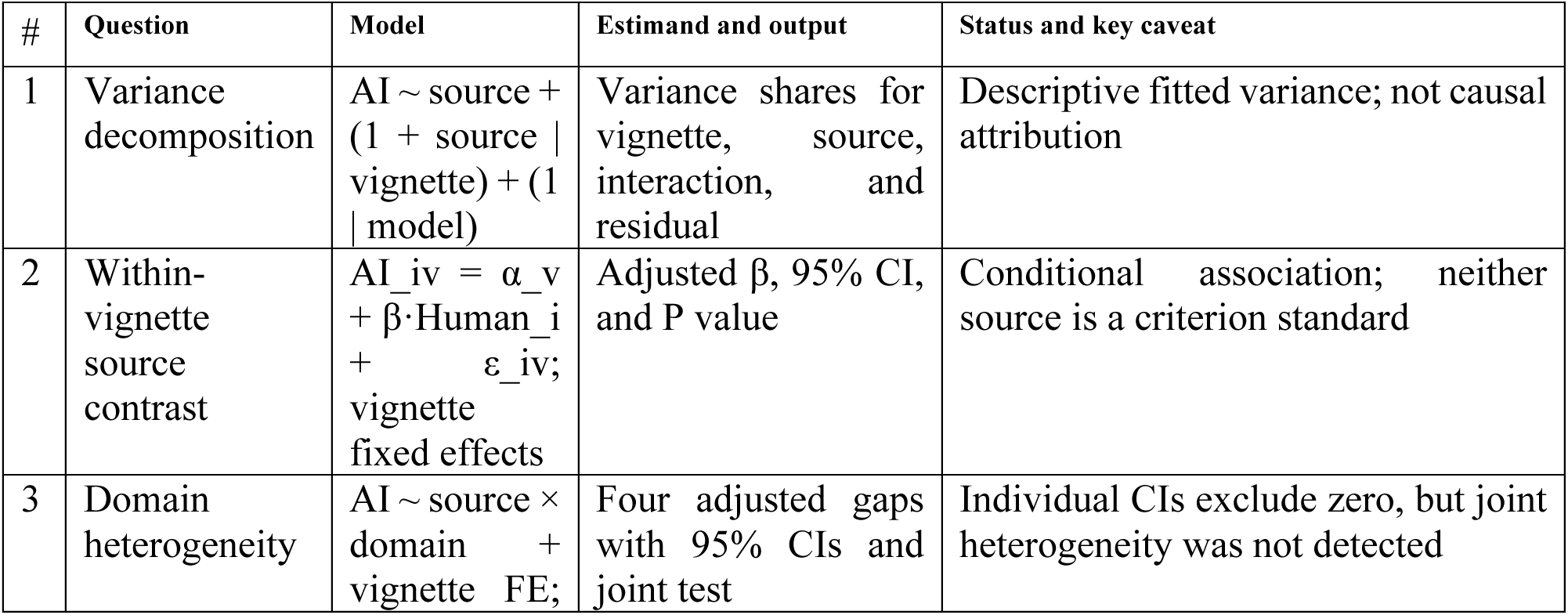
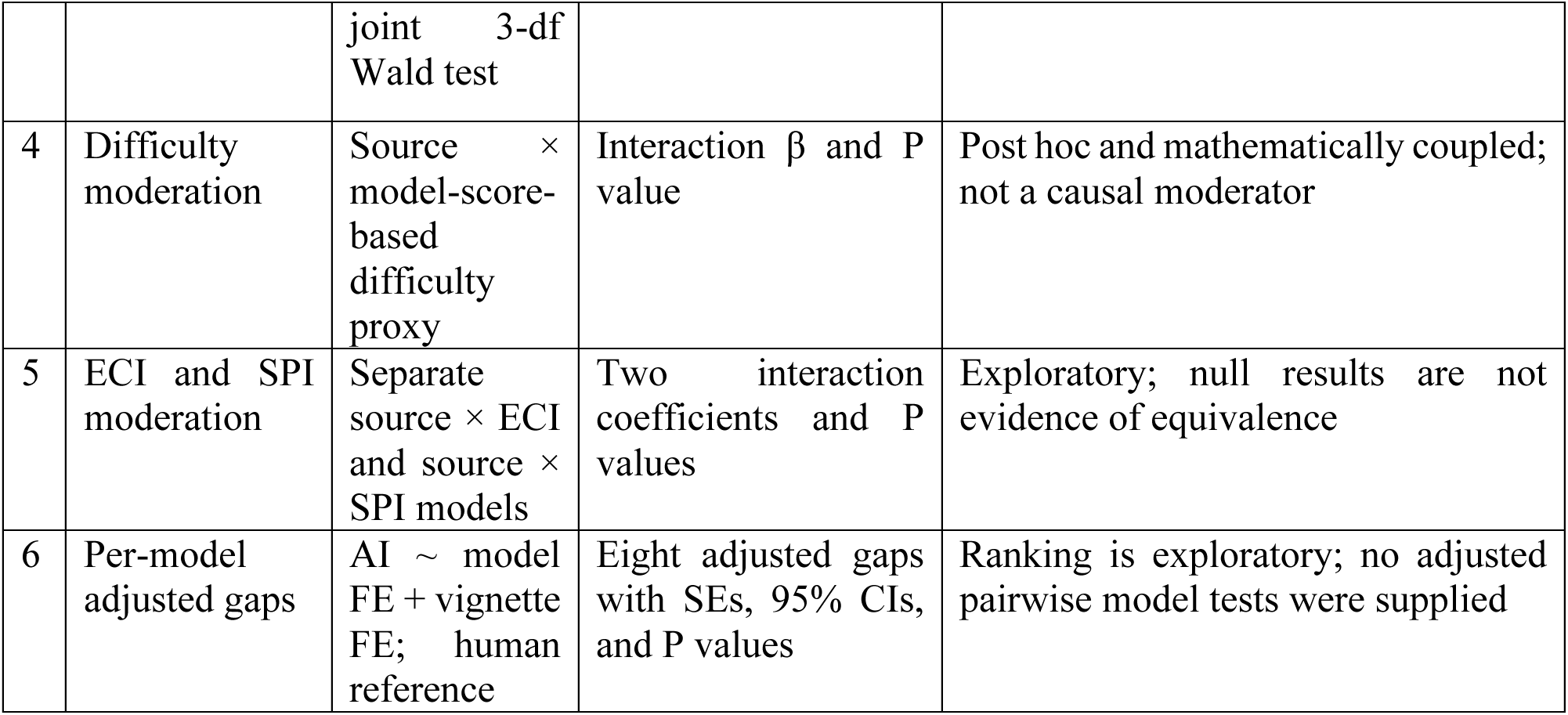
Exploratory explanatory models of rating differences.

**Table 9.** Human and model Autonomy Index on the harmonized scale, 50 comparison vignettes.

| **Metric** | **Value** |
| --- | --- |
| **Composite Autonomy Index** |  |
| Human mean | 50.2 |
| Model mean | 39.2 |
| Paired difference, human minus model | +11.0 (95% CI 6.3 to 15.8; $t = 4.64$, $df\ 49$, $P = 2.6 \times 10^{-5}$; $d_z = 0.66$) |
| **Domain difference, human minus model** |  |
| Factual Understanding; Intentional Action | +13.7; +12.3 |
| Values Awareness; Rational Deliberation | +10.4; +7.9 |
| Joint test of domain by source | Wald $\chi^2(3) = 2.27$, $P = 0.52$ |
| **Interpretive bands** |  |
| Strong (index $\geq 80$), human and model | 8% (4 of 50) and 8% (4 of 50) |
| Adequate (60 to 80), human and model | 30% (15 of 50) and 12% (6 of 50) |
| Reassess (index $< 60$), human and model | 62% (31 of 50) and 80% (40 of 50) |
| Same band assigned by both sources | 72% (36 of 50) |

**Table 10.**
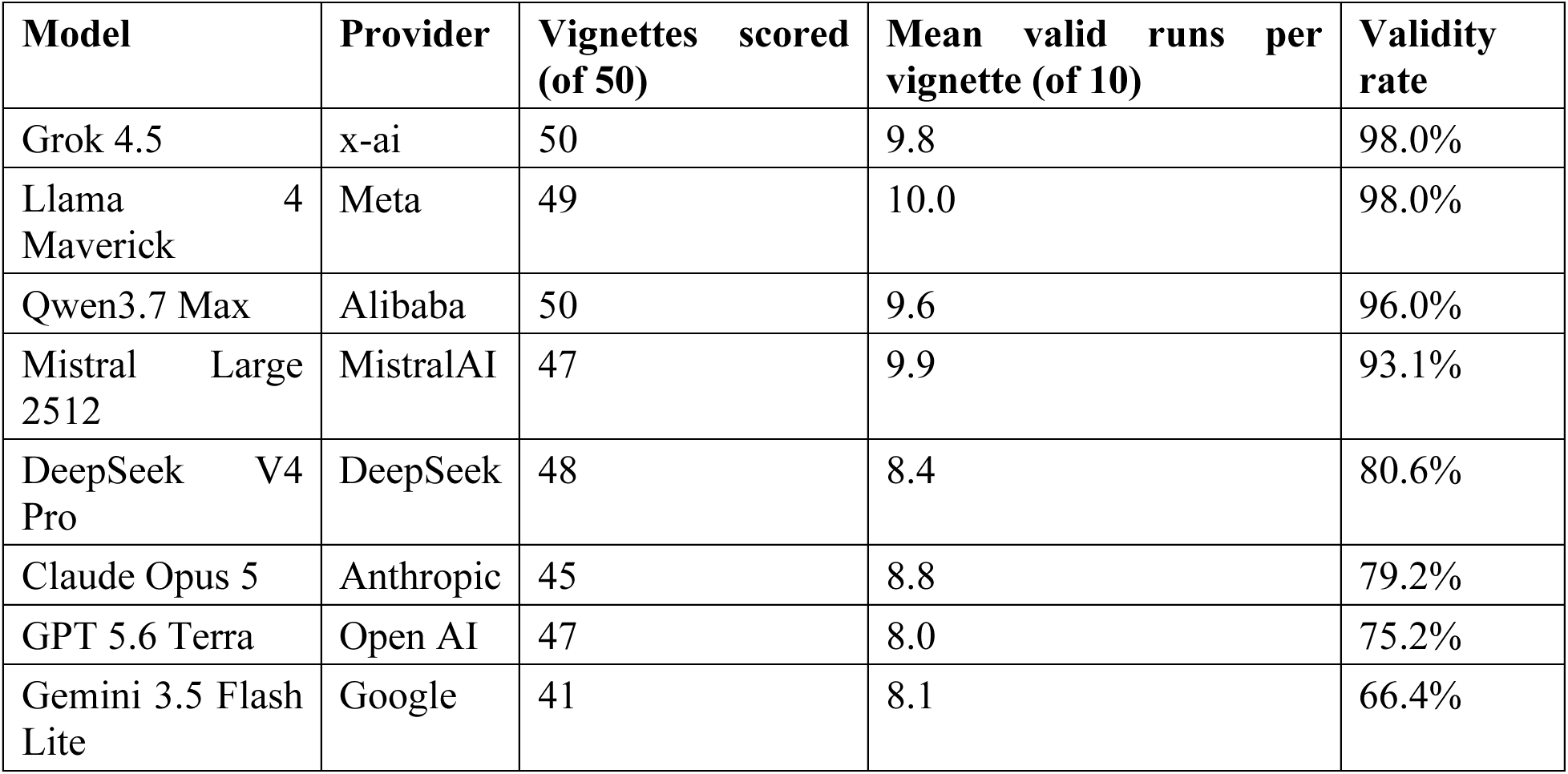
Model coverage and validity, July 2026 run, 50 comparison vignettes at ten runs per model per vignette.

The statistical analysis has three descriptive layers: central tendency and association, distributional divergence, and pluralistic coverage. Coverage analyses ask whether model scores fall within an operationally defined human range; they do not establish that any score in that range is correct. Table 7 summarizes the methods and interpretation guards.

Exploratory adjusted models estimate source contrasts after accounting for vignette and describe possible effect modification. Because source assignment was observational and case selection depended on model scores, the coefficients are not causal effects even when expressed as within-vignette contrasts. Table 8 summarizes the fitted models, reported outputs, and interpretation limits.

### 6.4 Human-rating completion

Of 275 assigned human-rating slots, 175 were returned; 173 ratings met the domain-completeness rule and entered analysis. Per-vignette counts ranged from 1 to 6. Assignment was round robin, but completion was not randomized, and reviewer characteristics were not linked to submission records.

The current analysis does not use inverse-probability weighting for noncompletion. Consequently, the human reference is conditional on observed submissions. Potential informative completion is treated as a limitation, and future validation should link assignment, reviewer characteristics, and completion so that response models can be estimated and reported.

## 7 Reporting conventions

Results are reported for the composite, four domains, ECI, SPI, and relevant interpretive flags. The paired 50-vignette estimate is the primary source comparison; mixed-model, quintile, distributional, coverage, and moderation analyses are secondary or exploratory. The composite is not interpreted without the domain profile.

Uncertainty was matched to each estimand. The primary paired mean difference uses the paired t distribution across 50 vignette-level differences; adjusted-model confidence intervals use model-based standard errors; and Bland–Altman limits use the mean difference ±1.96 standard deviations of the paired differences. Descriptive figures identify any standard error or other interval explicitly. No bootstrap interval is used for a primary inferential claim.

Interpretive bands are:

- AIx ≥ 80, strong evidence of autonomous agency
- AIx 60 to 79, adequate; additional supports may optimize the decision
- AIx < 60, autonomy not fully demonstrated; further support and reassessment indicated

Bands are interpretive aids and not decision rules. They are reported jointly with ECI and SPI so that a low score arising under high constraint or low support is read as an indication for environmental change rather than as a conclusion about the individual.

## Results

Results below compare human ratings with the July 2026 model generation on the 50-vignette comparison set, which is the primary comparison specified in section 5.2. Figures 4 to 11 accompany the text.

**Figure 4.**
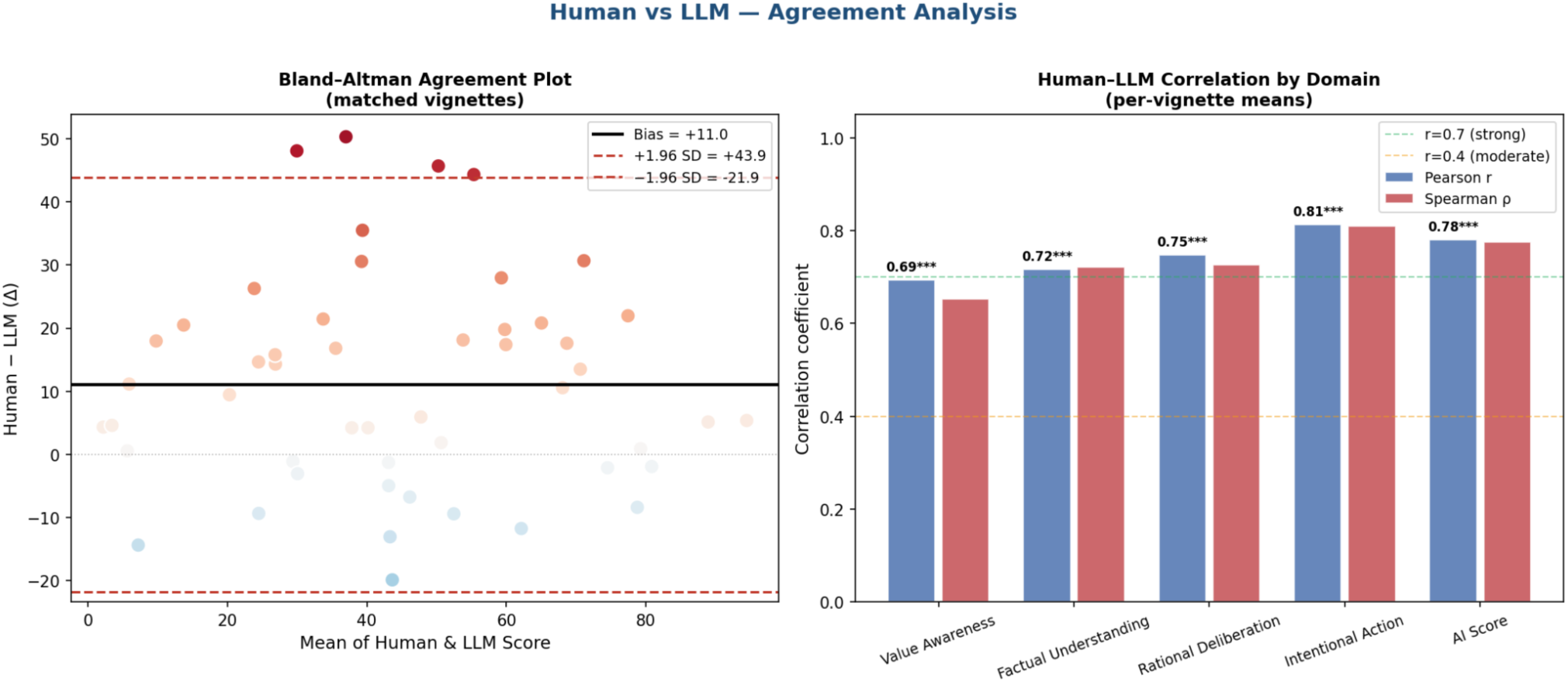
Agreement analysis on the 50 paired vignette-level source means. Left: Bland–Altman plot with mean human-minus-model bias of 11.0 points and 95% limits of agreement from −21.9 to 43.9. Right: Pearson and Spearman associations for the composite and four domains. Correlation describes ordering; the Bland–Altman panel shows that the sources are not interchangeable at the case level.

**Figure 5.**
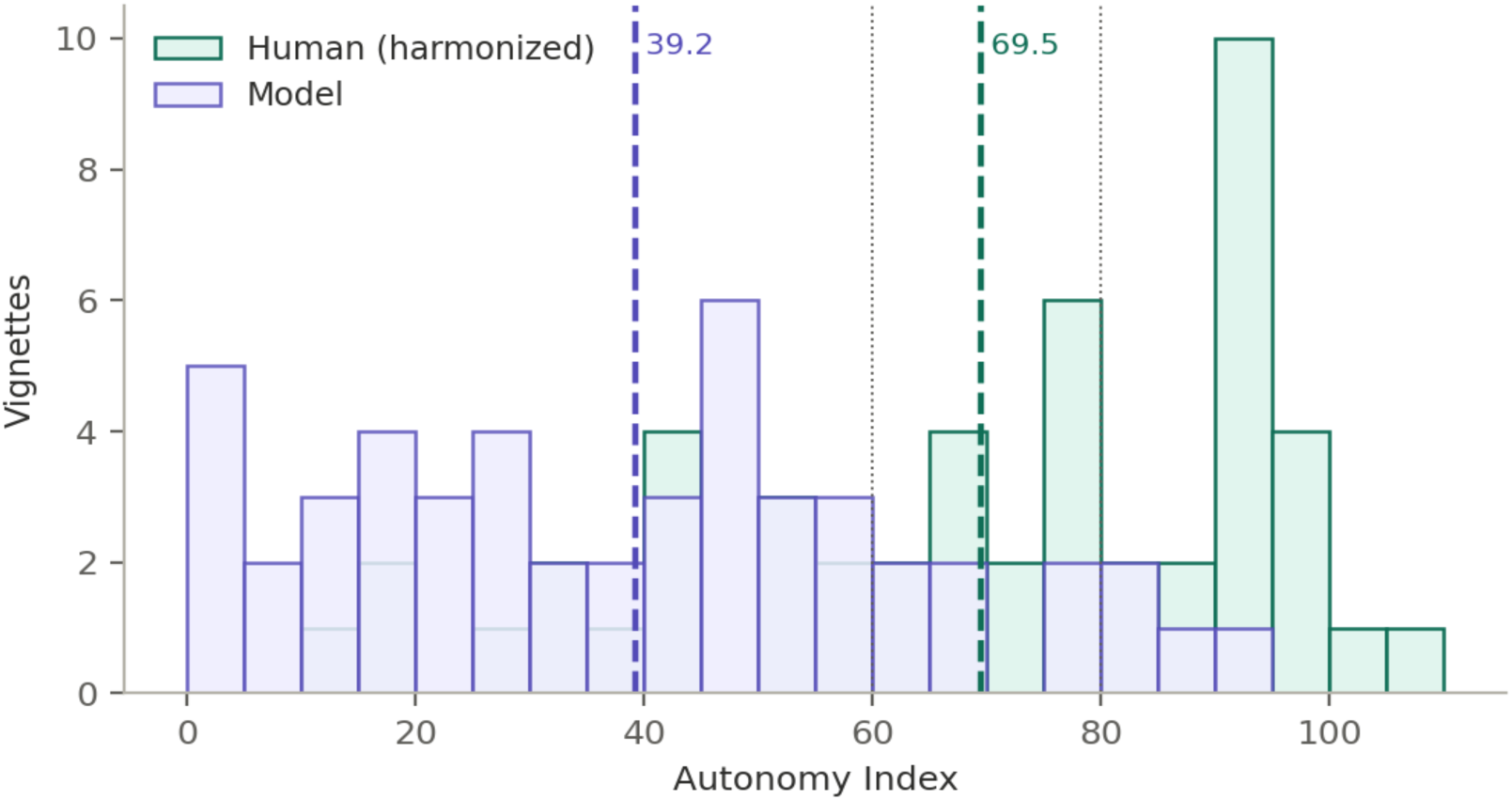
Distribution of the composite Autonomy Index for human reviewers and models across the 50 vignettes on the harmonized scale. Dotted vertical lines mark the interpretive band boundaries at 60 and 80. The two distributions have almost identical spread and are displaced, which is the pattern that rank agreement conceals and a level comparison exposes.

**Figure 6.**
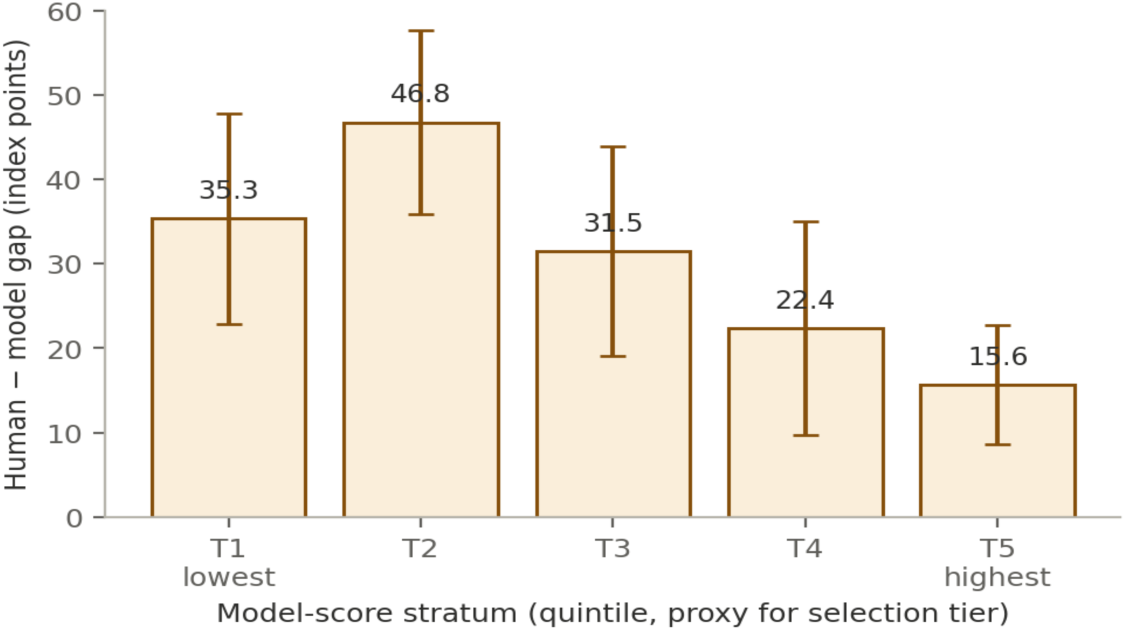
Human–model difference by quintile of the model composite, with 95% confidence intervals. Quintiles approximate the score tiers used for selection. The difference is largest where models assign the least autonomy and narrows as model-assigned autonomy rises.

**Figure 7.**
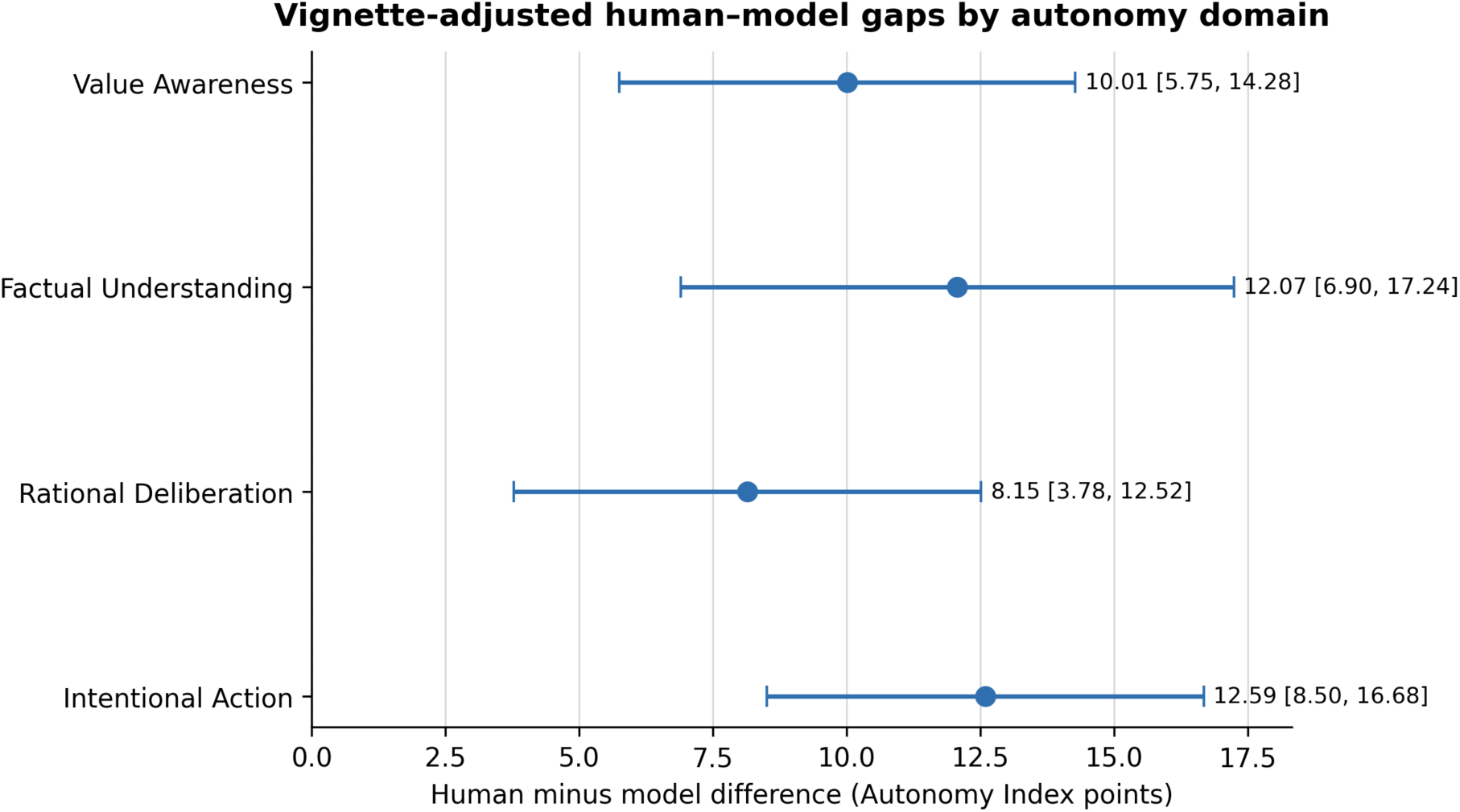
Vignette-adjusted human-minus-model differences by autonomy domain, with 95% confidence intervals. Every interval excludes zero, but the joint source-by-domain test did not detect heterogeneity.

**Figure 8.**
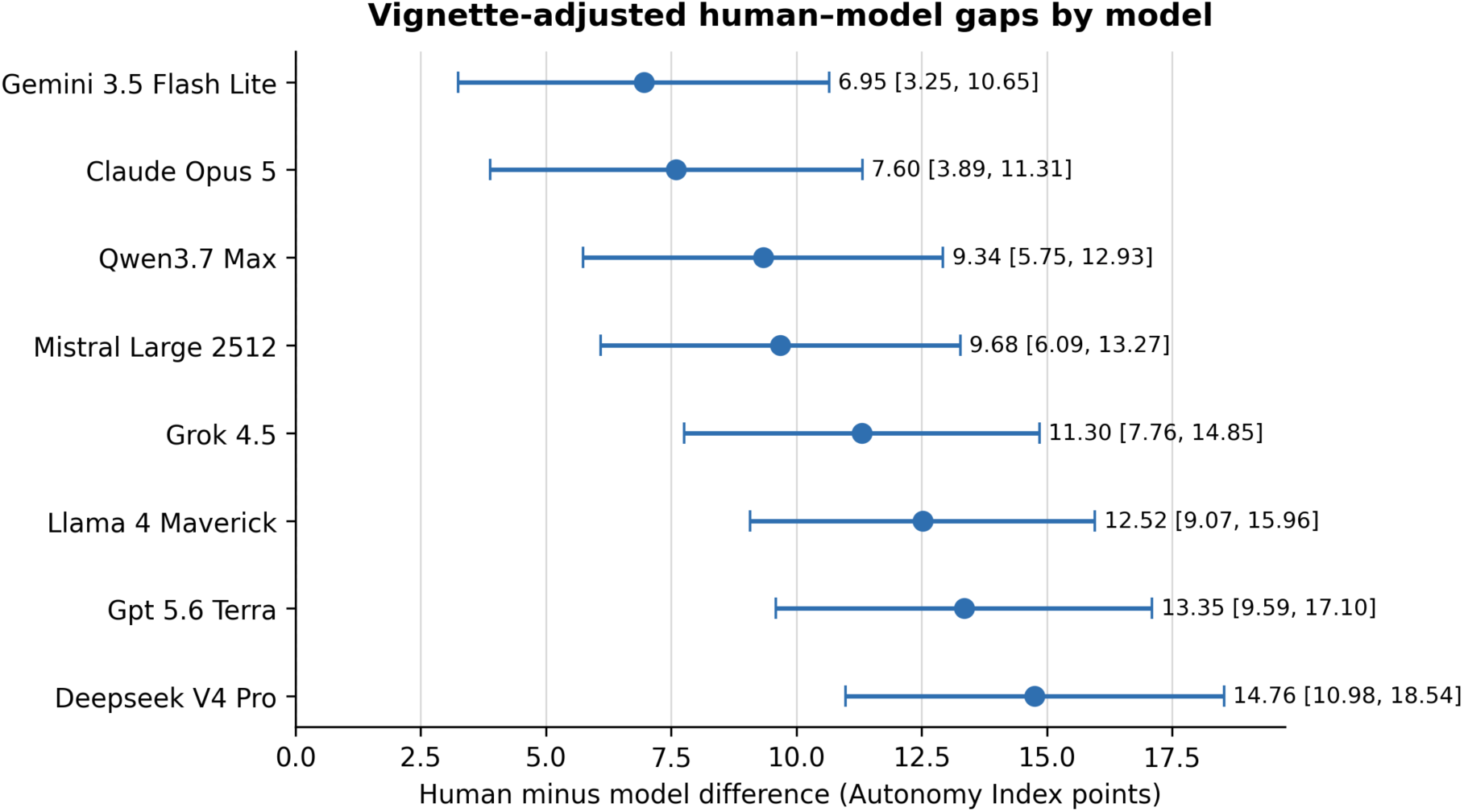
Vignette-adjusted human–model gap by model, with 95% confidence intervals, on the harmonized scale. Every interval excludes zero, and the intervals for the extreme models overlap only slightly.

**Figure 9.**
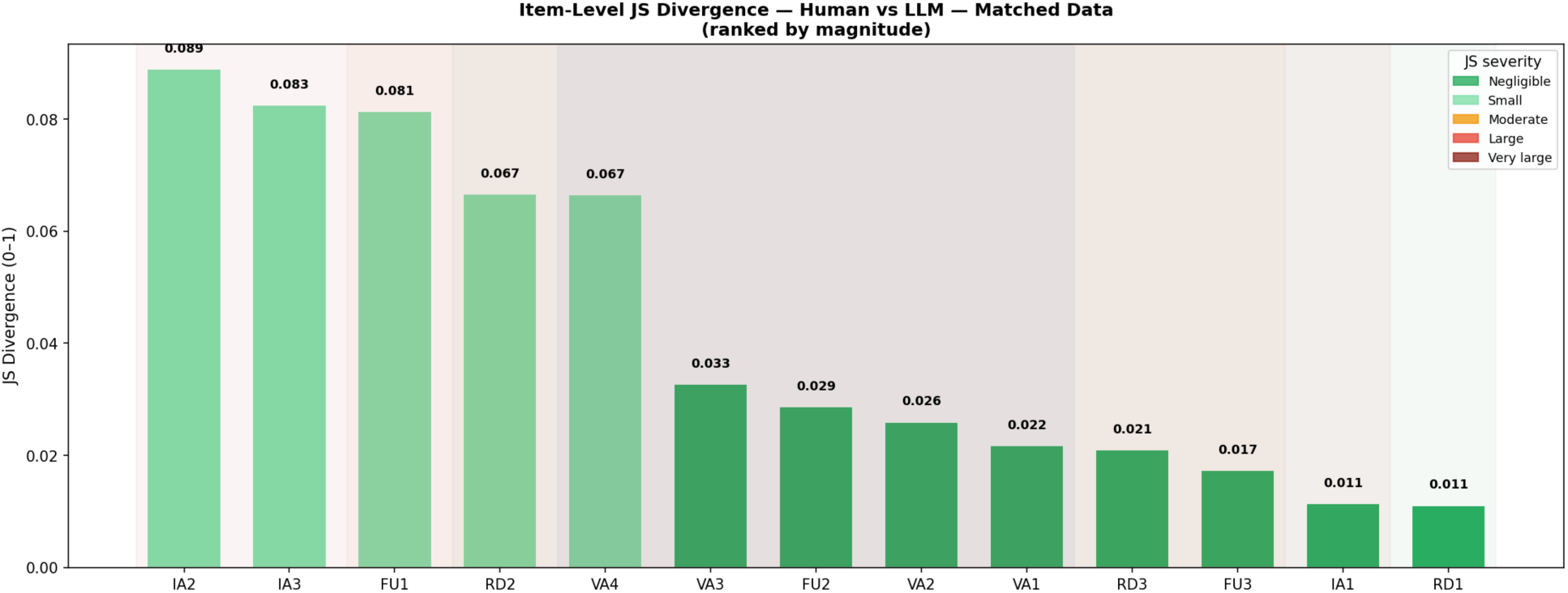
Jensen–Shannon divergence between human and model response distributions for each of the thirteen instrument items. Divergence is small or negligible at every item; the composite difference accumulates from consistent small shifts rather than from a few divergent items.

**Figure 10.**
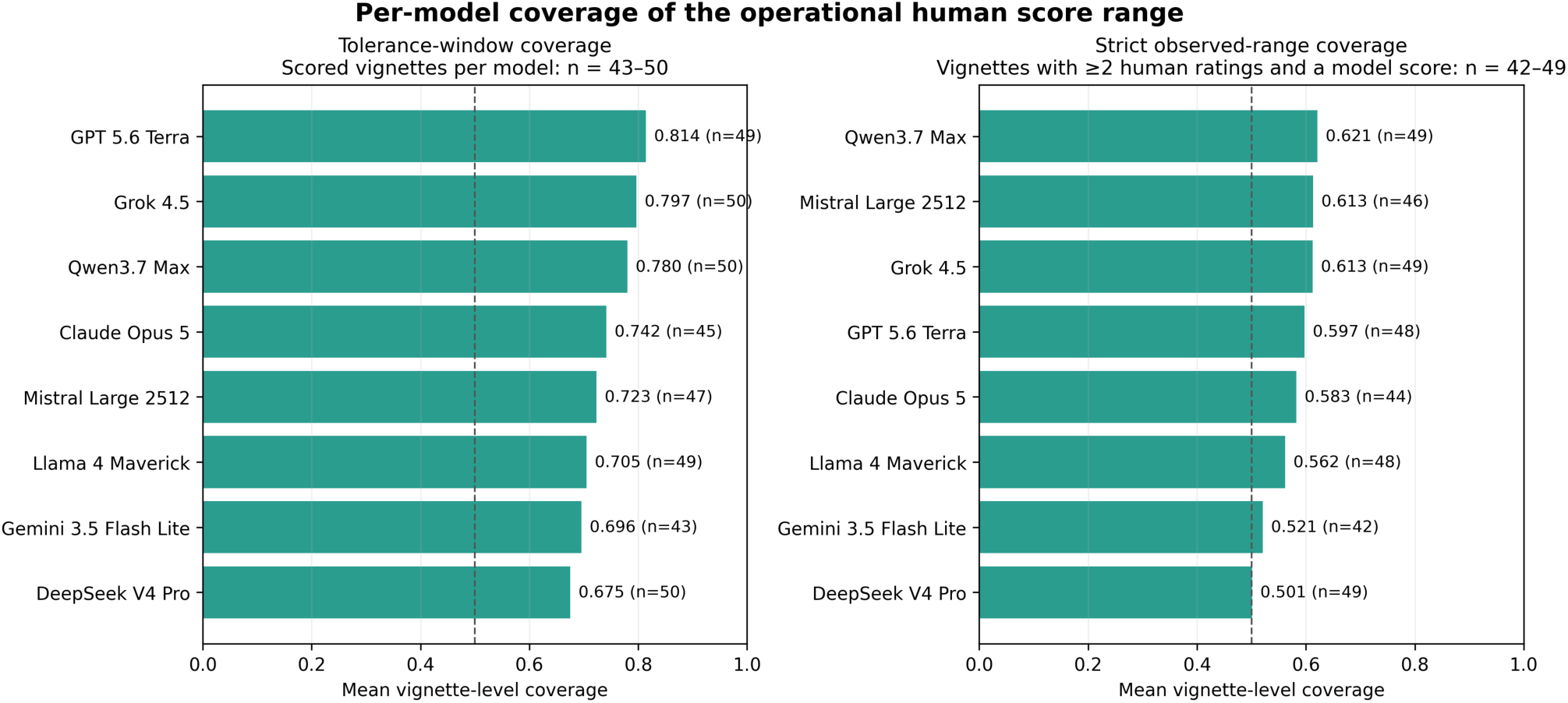
Mean vignette-level coverage of the operational human score range by model. The tolerance-window analysis includes 43 to 50 scored vignettes per model; the strict observed-range analysis requires at least two human ratings and includes 42 to 49 vignettes per model. Labels report the coverage estimate and the model-specific vignette denominator. The dashed 0.50 line is a descriptive reference, not a validated performance threshold.

**Figure 11.**
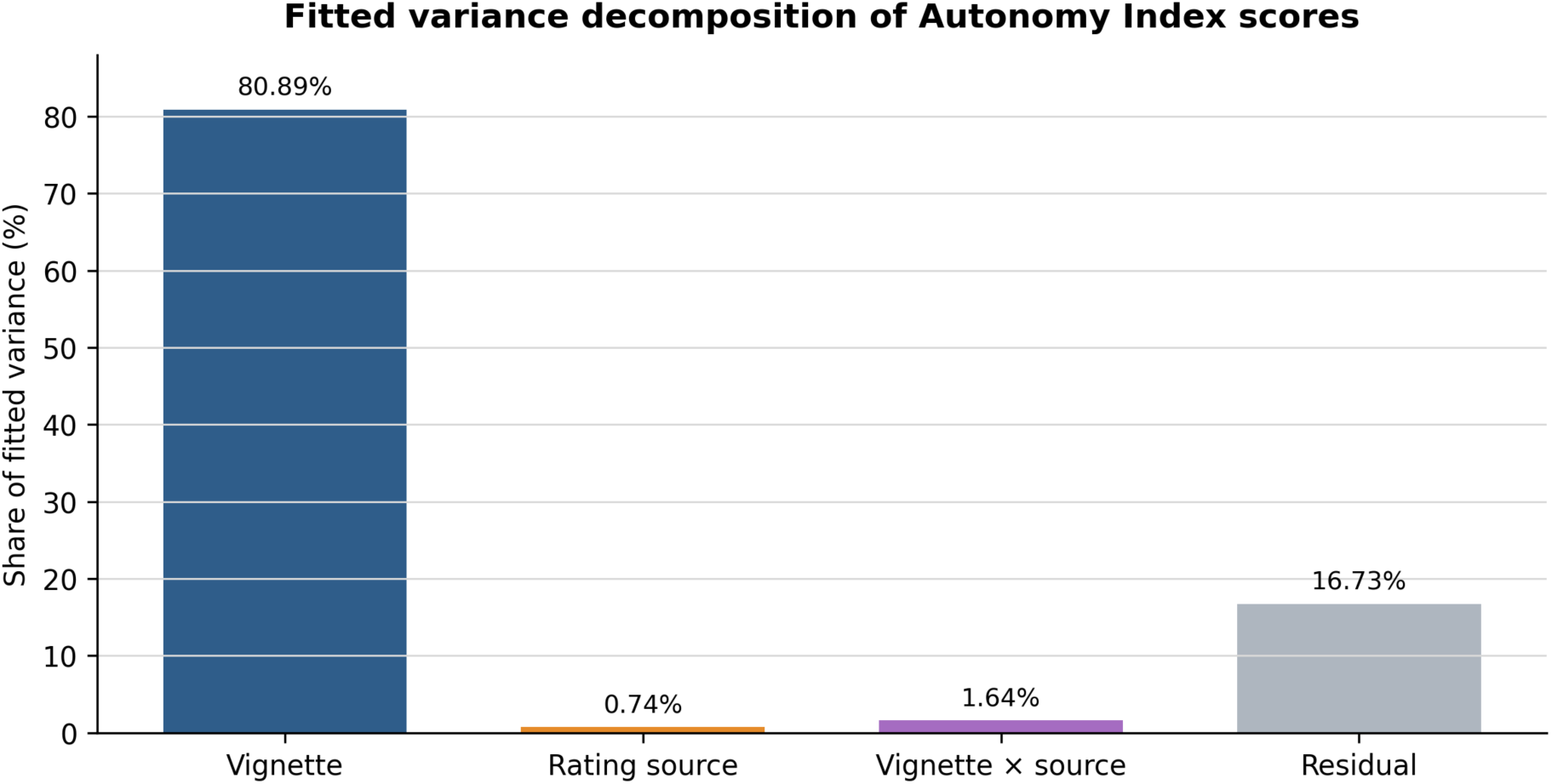
Fitted variance decomposition of Autonomy Index scores into vignette, rating-source, vignette-by-source, and residual components. The components describe the fitted comparison data and should not be interpreted as causal attribution.

### 8.1 The analyzed dataset and the response-scale harmonization

The analyzed file contains 173 human ratings across the 50 vignettes, together with 3,431 valid model scores from eight models scored in July 2026, each model run approximately ten times per vignette. Ratings per vignette range from 1 to 6 (median 3, mean 3.46, harmonic mean 3.09), with seven vignettes below the three-rating target and one vignette rated only once.

#### Response-scale quality control

Stored human response codes 1 through 5 were mapped to the intended ordinal scores 0 through 4; code 6 represented not applicable and was excluded from the relevant domain mean. Item scores were then normalized to 0 to 100 as specified in section 4.1. Range and logic checks confirmed that all analyzed domain and composite scores fell within 0 to 100.

### 8.2 Humans and models order cases alike

Across the 50 selected vignettes, human and model composites were strongly associated (Pearson r = 0.781, P = 2.3 × 10⁻¹¹; Spearman ρ = 0.775, P = 3.8 × 10⁻¹¹). Weighting vignettes by the number of human ratings gave Pearson r = 0.763. Domain-level Pearson correlations were 0.694 for Values Awareness, 0.717 for Factual Understanding, 0.747 for Rational Deliberation, and 0.813 for Intentional Action.

These coefficients indicate similar ordering in this comparison set, but they do not imply agreement in score level or clinical interchangeability. A disattenuated correlation was not calculated because the human-reference reliability model yielded a boundary estimate.

### 8.3 Models assign systematically less autonomy

The human mean was 50.2 and the model mean 39.2, a paired human-minus-model difference of 11.0 index points (95% CI, 6.3 to 15.8; t = 4.64; df = 49; P = 2.6 × 10⁻⁵; d_z = 0.66). The adjusted within-vignette source contrast was 11.32 points (95% CI, 7.64 to 15.00; P < 0.001). Human vignette means exceeded model means on 36 of 50 vignettes.

Bland–Altman bias was 11.0 points, with 95% limits of agreement from −21.9 to 43.9. Thus, the mean difference coexisted with wide case-level variation: for an individual vignette the model mean could exceed the human mean or fall substantially below it. These results do not support interchangeable use of the two sources for case-level scoring.

#### Exploratory variation across the score range

Grouping vignettes by quintile of the July model composite gave human-minus-model differences of 16.9, 19.5, 9.4, 5.9, and 3.3 points from the lowest to highest quintile. The correlation between model score and paired difference was −0.33 (P = 0.018).

This pattern may reflect variation in source differences, but it cannot distinguish that explanation from mathematical coupling, regression to the mean, or the model-based selection of vignettes. It is therefore hypothesis-generating and should be tested in a prospectively sampled set using an independently defined difficulty measure.

### 8.4 The difference is present in all four domains

Paired domain differences were 13.7 points for Factual Understanding, 12.3 for Intentional Action, 10.4 for Values Awareness, and 7.9 for Rational Deliberation. Corresponding vignette-adjusted gaps were 12.07 points for Factual Understanding (95% CI, 6.90 to 17.24), 12.59 for Intentional Action (8.50 to 16.68), 10.01 for Values Awareness (5.75 to 14.28), and 8.15 for Rational Deliberation (3.78 to 12.52). Although each interval excluded zero, the joint source-by-domain test did not detect heterogeneity (Wald χ²[3] = 2.27, P = 0.52). The observed ordering should therefore not be interpreted as evidence that the source difference is larger in one domain than another.

The domain estimates are compatible with a broadly distributed level difference rather than a difference isolated to one component. Because the analysis was not powered to establish equivalence across domains, the null interaction also does not prove that the four gaps are identical.

### 8.5 The difference is present for every model, but not equally

Vignette-adjusted gaps estimated by the mixed model range from 7.0 points to 14.8 points, with every confidence interval excluding zero. In order they are Gemini 3.5 Flash Lite 7.0 (3.3 to 10.6), Claude Opus 5 7.6 (3.9 to 11.3), Qwen3.7 Max 9.3 (5.7 to 12.9), Mistral Large 2512 9.7 (6.1 to 13.3), Grok 4.5 11.3 (7.8 to 14.8), Llama 4 Maverick 12.5 (9.1 to 16.0), GPT 5.6 Terra 13.3 (9.6 to 17.1) and DeepSeek V4 Pro 14.8 (11.0 to 18.5).

Adjusted human-minus-model gaps were positive for all eight models and ranged from 7.0 to 14.8 points. Because each model had incomplete and unequal vignette coverage, between-model ordering is exploratory; the estimates do not establish a common training or alignment mechanism. Model choice changed the magnitude of divergence in this sample but did not eliminate the positive mean gap.

### 8.6 Distributional divergence

Jensen–Shannon divergence^28^ between the human and model score distributions is 0.066 for the composite, which the thresholds used in the analysis classify as small. It rises across the domains, from 0.082 for Values Awareness (small) through 0.118 for Factual Understanding and 0.157 for Rational Deliberation (both moderate) to 0.212 for Intentional Action (large). The corresponding Jensen–Shannon distances, which are a proper metric on the unit interval, run from 0.257 for the composite to 0.460 for Intentional Action.

Kullback–Leibler divergence^29^ is close to symmetric at the composite level, 0.247 in one direction against 0.167 in the other, but becomes strongly asymmetric within domains: 2.61 against 0.32 for Factual Understanding, 2.73 against 0.46 for Rational Deliberation, and 4.35 against 0.55 for Intentional Action. The direction of the asymmetry is that human ratings visit score regions the models enter rarely rather than the reverse, and it is concentrated in the domains carrying the largest mean differences.

At item level the divergences are small throughout. Five of the thirteen items reach the small band — IA2 (plan-fulness, 0.089), IA3 (follow-through feasibility, 0.083), FU1 (key facts recall, 0.081), RD2 (trade-off reasoning, 0.067) and VA4 (appreciation, 0.067) — and the remaining eight are negligible, the smallest being RD1 (coherence, 0.011) and IA1 (intention strength, 0.011). No single item diverges enough to account for the composite difference, which is consistent with the item-level tests reported in section 8.12.

### 8.7 Models usually fall inside the range of human judgment

Under the prespecified tolerance-window definition, the mean vignette-level coverage was 0.741 across the 50 shared vignettes (median, 0.949). Thirty-eight of 50 vignettes had coverage of at least 0.50 and one had none. Per-model mean tolerance coverage ranged from 0.675 to 0.814 and was based on 43 to 50 scored vignettes per model. Under the strict observed human minimum-to-maximum definition, which required at least two human ratings and a valid score from the model, per-model coverage ranged from 0.501 to 0.621 across 42 to 49 eligible vignettes per model.

Coverage is sensitive to the number of human ratings and to the chosen tolerance window. It describes overlap between two empirical score distributions, not the proportion of clinically correct model outputs or a population prevalence. The result therefore complements, but does not explain, the paired mean difference.

Diversity match distinguishes two different patterns. Across models, the entropy-based diversity-match component ranged from 0.841 to 0.947 relative to the human response spread. At item level, the LLM-to-human entropy ratio ranged from 0.850 for IA2 to 1.053 for VA2; eleven of thirteen items were within 10% of the human entropy, while VA4 (0.872) and IA2 (0.850) showed the clearest narrowing. These values indicate broadly similar dispersion but do not show that the distributions are centered in the same place. The equal-weight exploratory pluralistic score (one-half coverage and one-half diversity match) ranked Grok 4.5 highest at 0.872, followed by Qwen3.7 Max at 0.864 and GPT 5.6 Terra at 0.863; DeepSeek V4 Pro was lowest at 0.765. Because the one-half/one-half weighting is an analytic choice, the component values remain the primary descriptive results.

### 8.8 Variance decomposition and moderators

In the exploratory variance decomposition, vignette content accounted for 80.89% of fitted Autonomy Index variance, rating source for 0.74%, the vignette-by-source interaction for 1.64%, and residual variation for 16.73%. The adjusted within-vignette human-minus-model contrast was 11.32 points (95% CI, 7.64 to 15.00; P < 0.001). Variance shares and mean contrasts answer different questions: a small source variance component can coexist with a nonzero average source difference. These estimates describe the selected comparison data and are not causal variance attribution or causal effects of rater type.

Exploratory moderation. The source-by-difficulty interaction was negative (β = −0.2507, P = 0.0003), consistent with a narrower human–model gap as the model-score-based difficulty proxy increased. However, because that proxy was constructed from the analyzed model scores, the coefficient is mathematically coupled to the outcome and remains hypothesis-generating. Neither the ECI interaction (β = −0.0116, P = 0.8912) nor the SPI interaction (β = 0.0940, P = 0.1827) provided evidence of contextual moderation.

The null ECI and SPI interactions do not establish absence of moderation, and the study was not designed or powered as an equivalence analysis. The apparent difficulty interaction may reflect mathematical coupling, regression to the mean, or selection of the comparison set rather than genuine effect modification. No mechanistic or causal conclusion is drawn from these exploratory coefficients.

### 8.9 Where humans and models agree: the contextual modifiers

The two contextual modifiers behave very differently from the autonomy domains. Human and model External Constraint Index scores correlate at r = 0.82 (P = 4.5 × 10⁻¹³) across the 50 vignettes, and their means differ by 6.4 points on a 0 to 100 scale, 62.4 for humans against 68.8 for models, a difference that does not survive Holm correction across the fifteen scored quantities (adjusted P = 0.49). Support Provided Index scores correlate at r = 0.40 (P = 0.004) and differ by 3.0 points, 44.7 against 47.7, also not significant (adjusted P = 0.49). Both differences run opposite to the autonomy difference: models read slightly more constraint and slightly more support into the same cases while reading less agency into the person.

ECI and SPI provide useful comparators, but they are not formal negative controls: they measure different constructs and use different item structures from the thirteen-item autonomy composite. The non-significant mean differences therefore do not demonstrate that humans and models read every vignette circumstance identically.

The results suggest stronger source divergence for the autonomy composite than for the contextual modifiers in this sample. They do not identify where in the rating process that divergence arises.

### 8.10 Interpretive band concordance

Humans and models assign the same interpretive band on 36 of the 50 vignettes, 72 percent. The two sources agree exactly at the top of the scale, each placing four vignettes in the strong band above 80. They differ in the middle: models place 40 of 50 in the reassess band against 31 for humans, and 6 in the adequate band against 15. The disagreement is therefore concentrated at the 60-point boundary, where a case human reviewers read as adequate is read by models as warranting reassessment, and it is absent at the top of the scale.

Band concordance is a descriptive transformation of the same underlying scores and depends on provisional 60- and 80-point thresholds. It should not be interpreted as an independent validation result or as evidence about clinical classification accuracy.

### 8.11 Psychometric performance

For July model ratings, the variance-component analysis estimated vignette, model, and residual variances of 637.3, 7.0, and 116.8, respectively. Single-rating generalizability was 0.837, and aggregate reliability was 0.975 at the realized mean panel size of 7.7 models per vignette. The decision-study projection exceeded the 0.80 threshold with one model rating and increased with panel size. The analogous human model yielded a boundary solution: vignette variance was estimated at zero, reviewer variance was not estimable, and residual variance was 339.6. Human generalizability coefficients and decision-study projections were therefore not interpreted. The boundary result reflects the sparse, incomplete rating design and should not be read as evidence that human judgments contain no shared signal.

Internal consistency across the thirteen autonomy items was high for both sources. Ordinal alpha was 0.978 for July model ratings and 0.971 for human ratings; the corresponding Pearson-based values were 0.968 and 0.959. These estimates support coherent composite scoring in this sample, but they do not establish unidimensionality or show that the four conceptual domains are interchangeable.

Run-to-run self-consistency was generally high but varied by model. ICC(1) and within-vignette standard deviation were, respectively: Mistral Large 2512, 0.998 and 0.72 (47 vignettes; mean 9.9 valid runs); Claude Opus 5, 0.990 and 2.16 (45; 8.8); Gemini 3.5 Flash Lite, 0.974 and 3.95 (41; 8.1); Llama 4 Maverick, 0.955 and 3.81 (49; 10.0); GPT 5.6 Terra, 0.953 and 4.48 (47; 8.0); Grok 4.5, 0.943 and 4.46 (50; 9.8); Qwen3.7 Max, 0.912 and 6.98 (50; 9.6); and DeepSeek V4 Pro, 0.825 and 10.62 (48; 8.4). Thus, strong aggregate reliability across models did not imply equivalent repeatability for every individual model.

### 8.12 Item-level source differences

Item-level comparison with Holm correction across the fifteen scored quantities identified five differences that remained statistically significant: VA1 value clarity (human 2.59 versus model 2.18, adjusted P = 0.002), VA2 value stability (2.56 versus 2.10, adjusted P = 0.001), VA4 appreciation (1.61 versus 1.15, adjusted P = 0.035), IA2 plan-fulness (1.69 versus 1.07, adjusted P = 0.0004), and IA3 follow-through feasibility (1.99 versus 1.25, adjusted P < 0.001). The remaining eight autonomy items and both contextual modifiers did not remain significant after correction, although all thirteen autonomy item differences were in the same direction. The composite difference therefore reflects an accumulation of directionally consistent item-level shifts rather than a difference confined to a few items.

### 8.13 Model coverage, validity, and not-applicable responses

Section 5.2 specifies 10 runs per model per vignette, so the July run attempted 500 scorings per model across the 50 comparison vignettes and 4,000 in total. It returned 3,431 valid parsed responses, an overall validity rate of 85.8 percent, and that rate varies substantially by model. Table 10 reports coverage and validity for each.

Valid runs per vignette are averaged over the vignettes a model scored at least once, so the validity rate is the product of the two preceding columns divided by 500, the scorings attempted per model.

Two features of this table matter for interpretation. Coverage is incomplete for six of the eight models: Gemini 3.5 Flash Lite returned at least one valid score on 41 of the 50 vignettes, Claude Opus 5 on 45, and Mistral Large 2512 and GPT 5.6 Terra on 47 each. The per-model gaps in section 8.5 are therefore estimated on partly different vignette subsets, which the mixed model handles by conditioning on vignette rather than by imputation, but which no analysis can fully repair.

The second feature invites a reading that the data do not support. The model with the smallest gap, Gemini 3.5 Flash Lite, is also the model with the lowest validity, which suggests that differential attrition might explain the per-model ordering in section 8.5. Across the eight models the rank correlation between validity rate and vignette-adjusted gap is 0.26 (P = 0.53), and the two models with the largest gaps have validity rates of 75.2 and 80.6 percent, in the same range as the model with the smallest gap. Differential validity is a genuine limitation of the per-model comparison; it is not a sufficient explanation of it.

#### Not-applicable responses

The two sources differ sharply in how often they decline to score an item. Models recorded no not-applicable responses at all, on any item, for any vignette, giving an overall model rate of 0.00 percent. Human reviewers recorded not-applicable on 22.5 percent of items. The human rate varies by domain, from 19.9 percent for Values Awareness through 22.5 percent for Intentional Action and 30.4 percent for Rational Deliberation to 33.1 percent for Factual Understanding, and by item from 6.4 percent at VA1 (value clarity) to 38.4 percent at RD2 (trade-off reasoning). Neither contextual modifier drew a single not-applicable response from either source.

The absence of model not-applicable responses is a clear behavioral difference, but its cause and effect on composite scores cannot be identified here. Across only four domains, the rank correlation between human not-applicable rate and mean source gap was 0.40 (P = 0.60); that low-powered ecological comparison neither confirms nor rules out forced scoring as a contributor. A direct test would require rerunning models under experimentally varied instructions or scoring rules.

### 8.14 Landmark and higher-coverage vignette check

Nineteen vignettes had at least four human ratings. Nine had human-minus-model differences greater than 15 points: Vaccine Hesitancy (+17.4), Alzheimer consent (+22.0), Everything Be Done (+16.8), Too Little Too Late (+20.8), A Family Divided (+20.5), Maxine (+35.5), Genetic Testing (+19.8), Literacy (+17.6), and Distributive Justice (+44.3). No vignette in this subset had a difference below −15 points; the full observed range was −13.1 to +44.3. These case-level results are descriptive and remain conditional on the selected comparison set and the number of available human ratings.

## Discussion

Across this selected comparison set and common instrument, trained reviewers assigned autonomy scores averaging 11.0 points higher than eight general-purpose language models. The sources ordered vignettes similarly, but wide limits of agreement show that the average difference is not a reliable case-level correction.

The positive mean difference appeared across all models and domains, although model coverage was incomplete and the joint domain test did not detect heterogeneity. The adjusted models confirmed that the direction of the source contrast persisted after controlling for vignette, but they do not establish whether the pattern arises from training, alignment, prompt interpretation, differential use of not applicable, model attrition, human judgment, or another mechanism. The contextual-modifier and variance-decomposition results narrow some descriptive possibilities but do not identify a causal pathway.

The results support caution if these systems are used to structure autonomy or capacity discussions: a model score should not substitute for an interactive clinical assessment or be treated as a calibrated estimate of capacity. Human review remains necessary, not because this study proves that every lower model score is erroneous, but because human ratings are not ground truth, case-level differences are wide, and neither the instrument nor its interpretive bands has been clinically validated for decision making.

### 9.1 What the human ratings show about human judgment

Human ratings also warrant study as a measurement process. Reviewers assigned a mean composite of 50.2 and used nearly the full 0-to-100 range. In the exploratory decomposition, vignette content accounted for most fitted variance, but sparse and unbalanced ratings prevented a stable human reliability estimate. The findings therefore show differentiation across cases in the observed sample without establishing how reproducible a new reviewer panel would be.

The domain profile is flatter for reviewers than for models, though both order the domains the same way. Human domain means span 6.9 points, from 47.7 for Factual Understanding to 54.7 for Values Awareness, with Rational Deliberation and Intentional Action between them. Model means span 10.2 points across the same domains, from 34.0 to 44.3. Both sources rate Factual Understanding lowest and Values Awareness highest; models simply separate the domains further than reviewers do.

Two interpretations of the comparatively flat human domain profile are plausible, and this study cannot separate them. Reviewers may hold an integrated view of autonomy in which the components move together, or an overall impression of a case may propagate across the thirteen items as a halo effect. Distinguishing these possibilities would require a dedicated design, such as isolated domain scoring or manipulation of item order. The present internal-consistency estimates do not resolve that question.

### 9.2 Values in practice, and value in care

This work developed an instrument that breaks autonomy assessment into subcomponents and quantifies it. It made explicit which considerations a rater weighted and by how much. Applied to human reviewers, it externalized professional judgment that normally remains implicit, and when applied to LLMs, it compared the autonomy gap between humans and models.

This has a practical extension. Decisions about aggressive treatment, discharge planning, surrogate authority, and resource allocation carry both ethical and economic weight, and these dimensions are usually discussed in separate registers. A structured record of how autonomy was assessed at the point of decision makes it possible to ask whether decisions that honor patient agency align with or diverge from decisions that deliver high-value care, rather than assuming the two either coincide or conflict. The present study does not answer that question. It supplies the measurement layer that would let it be asked with evidence rather than assertion, and the domain-level profile is what makes that possible, since the composite alone would collapse exactly the distinctions that matter.

### 9.3 What this study does not establish

The observed 11-point mean difference is estimated for this selected set, but the design cannot establish which source is better calibrated. Human reviewers constitute a reference standard rather than ground truth, and the Autonomy Index itself requires external validation against independent clinical and patient-centered criteria.

Two design changes would address the largest uncertainties: denser, more balanced human ratings to estimate reference reliability, and a prospectively sampled vignette set independent of model scores. A preregistered replication should also specify the primary estimand, missingness strategy, coverage tolerance, and multiplicity families before analysis.

## 10 Limitations

The human–model difference is a mean over cases with wide case-level variability. The 95 percent limits of agreement span − 21.9 to 43.9 index points, so on an individual vignette a model score may exceed the human mean by twenty points or fall short of it by forty. The aggregate difference is well estimated; the per-case difference is not. No statement in this study should be read as a prediction about a particular case.

The analyzed dataset comprises 173 of the 175 ratings submitted at the analysis freeze, the difference being ratings excluded under the domain-completeness rule of section 3.5. Per-vignette weighting and the reliability estimates that depend on rating counts are conditional on that distribution. The analytic dataset was frozen on June 26, 2026; submissions received after that date were not included.

Vignettes are static written cases and cannot capture the interactive, temporally extended character of an actual capacity assessment, in which a clinician can probe, re-explain, and observe change. Scores therefore describe autonomy as depicted rather than autonomy as it would be established at the bedside.

Anchors are generic rather than item-specific, which places interpretive weight on the rater and is a plausible contributor to observed between-rater variance.

Two item pairs contain overlapping wording. RD1 asks whether reasoning is coherent and consistent while RD3 asks whether the decision is consistent with stated values and understanding. VA1 asks how clearly the subject articulates values relevant to the decision while VA3 asks whether the subject shows awareness of how their values relate to the decision. These pairs are candidates for targeted inter-item analysis and possible rewording in a future validation study; inter-item correlations for these pairs were not part of the current reported analysis.

The rater-by-vignette design is incomplete, so reviewer effects are estimated from partially overlapping data. The boundary solution in the human variance model precluded interpretable human reliability coefficients and underscores the need for denser, more balanced reviewer allocation.

Ratings are unevenly distributed across reviewers. The five most active reviewers contributed 64 of 173 ratings, 37.0 percent of the corpus, so the human variance model depends disproportionately on a small number of individuals. This imbalance is an additional reason not to interpret the boundary reliability estimate as a substantive finding.

The comparison set was stratified on model-derived scores rather than sampled at random, and this is the most consequential limitation of the design. Because the stratifying variable is the quantity under comparison, the comparison set is not a sample of the corpus in any respect that bears on the primary analysis. Descriptive distributions therefore characterize the comparison set alone: the equal allocation across five score tiers guarantees a flatter distribution of model scores than the corpus contains, so no statement about the prevalence of low-autonomy cases can be read off these 50 vignettes.

This selection cannot be fully repaired analytically. Quintile, moderation, and coverage results remain conditional on the selected set, and oversampling high-disagreement vignettes directly affects agreement measures. The difficulty analysis has an additional limitation because its predictor was derived from the same model scores used to define the human–model gap, creating mathematical coupling. A prospective replication should use a randomly drawn or consecutively assembled sample and an independently defined difficulty measure.

Selection also favored vignettes with sufficient narrative content to support scoring, which selects toward cases on which raters are less likely to record not applicable. The human–model contrast in not-applicable rates reported in section 8.13 is therefore a conservative estimate of what a randomly sampled corpus would show.

Three widely taught landmark cases were deliberately included. Reviewers may have recalled published commentary rather than rating only the vignette text. In the broader higher-coverage check, 19 vignettes had at least four human ratings; 9 showed human-minus-model gaps greater than 15 points and none showed a gap below −15 points. These descriptive results do not isolate recall effects in the three landmark cases.

The reviewer sample is a convenience sample rather than a probability sample, and its composition is concentrated in two respects. Of the 36 registrants who stated a specialty, 17 practice consultation-liaison psychiatry. That concentration cuts both ways. Assessment of decisional capacity is routine consultation work in that specialty, so the reference standard is weighted toward reviewers who perform the underlying judgment regularly, which strengthens its claim to expertise. But it also means the standard reflects the norms of one specialty more than the norms of clinicians generally, and agreement between models and, say, primary care physicians or intensivists cannot be inferred from these results.

Reviewer characteristics were collected at registration and are not linked to submission records, so contributing and non-contributing reviewers cannot be compared on role, experience, or specialty. This matters more than it would in a study with near-complete participation: 15 of the 45 reviewers holding assigned blocks submitted no ratings, and if that exit was associated with seniority, specialty, or available time, the realized reference standard differs from the sample described in Table 4 in a way the present data cannot detect. Linking registration characteristics to submission records in a future round would close this gap at negligible cost.

Reviewers were told at consent that their ratings would be compared with ratings produced by large language models. Disclosure was the right ethical choice, but it means the human ratings were not produced blind to the study question. A reviewer who knows a machine comparison is coming may deliberate more carefully than they would in routine practice, or may adjust scores against an imagined machine standard in either direction. The magnitude and direction of any such effect are not estimable from this design, since there is no undisclosed comparison group. Human ratings should therefore be read as expert judgments made under known comparison rather than as a naturalistic sample of clinical reasoning, and a replication with disclosure withheld until debriefing would be required to bound the effect.

Human reviewers were drawn from a clinically and bioethics trained population, and their judgments constitute a reference standard, not ground truth. Where models diverge from human ratings, the finding is divergence from trained human judgment, which is the relevant benchmark for a decision-support context but is not equivalent to error.

## 11 Conclusions

In the deliberately selected 50-vignette comparison set, trained reviewers assigned autonomy scores averaging 11.0 points higher than eight general-purpose language models, while the two sources ranked cases similarly. Positive mean gaps appeared in all four domains and for every model, but model coverage was incomplete and the joint domain test did not detect heterogeneity. Wide limits of agreement show that the mean difference cannot be applied as a case-level correction.

The study demonstrates measurable divergence from a trained-human reference; it does not establish model error, human correctness, a causal mechanism, or clinical validity. Model-based case selection, sparse human ratings, convenience sampling, and the absence of an independent outcome limit generalization. Until the instrument and model behavior are prospectively validated, model scores should be used only as research outputs or prompts for structured human deliberation, not as substitutes for capacity assessment or patient-centered clinical judgment.

## Declarations

### Ethics approval and consent to participate

Human data collection was reviewed and approved by the Harvard Longwood Campus Institutional Review Board under protocol ***IRB26-0146***. All participants provided documented electronic informed consent before accessing any study material; consent was recorded by affirmative selection on a dedicated consent screen and archived as a timestamped record separate from response data. No identifiable patient data were used at any point. All clinical vignettes were derived from published case reports and public-domain clinical material and contain no direct or indirect patient identifiers. The study was conducted in accordance with the Declaration of Helsinki and the Belmont Report.

### Consent for publication

Not applicable. The manuscript contains no identifiable individual-level information.

### Availability of data and materials

De-identified vignette-level and item-level rating data, together with the full instrument as presented to both human reviewers and models, are archived at Zenodo (https://doi.org/10.5281/zenodo.22802309) and mirrored at https://github.com/taposh/autonomyindex. The rating platform at https://medethics.org/ hosts the same materials but requires free registration. Individual reviewer identifiers and registration data are not shared, as participants did not consent to the release of individual-level records.

### Code availability

Scoring, aggregation, and analysis code, including the per-model configuration used for every API call, is archived at Zenodo (https://doi.org/10.5281/zenodo.22802309) and available at https://github.com/taposh/autonomyindex under the MIT License.

### Author contributions

T.D.R.: conceptualization, methodology, software, formal analysis, investigation, data curation, visualization, writing — original draft. R.W.B.: conceptualization, supervision, review and editing. Both authors read and approved the final manuscript.

### Use of generative artificial intelligence

Large language models were the object of study in this work. Model identifiers, providers, prompts, routing, and available configuration details are reported in the manuscript and accompanying code repository. We also used generative AI tools for code drafting and debugging, language editing, and figure development. The authors reviewed and verified all analyses and manuscript content and take responsibility for the accuracy, integrity, and originality of the work. No AI tool is listed as an author.

### Pre-registration

The analysis plan described in Section 6.3 was not pre-registered. The primary estimand, domain structure, tolerance-window definition, and multiplicity family were specified before analysis of the July comparison dataset, but the analysis plan was not publicly preregistered. All remaining analyses should be considered exploratory.

## Data Availability

De-identified vignette-level and item-level rating data, together with the full instrument as presented to both human reviewers and models and the scoring and analysis code, are archived at Zenodo (https://doi.org/10.5281/zenodo.22802309) and mirrored at (https://github.com/taposh/autonomyindex. The rating platform at https://medethics.org/ hosts the same materials but requires free registration. Individual reviewer identifiers and registration data are not shared, as participants did not consent to the release of individual-levelrecords.

https://github.com/taposh/autonomyindex

## Acknowledgements

The authors would like to thank Kelsey Flynn, Dr. Ana Lewis, Kristina Larson, Dr. Ed Hundert, and other faculty from Harvard Medical School and the Center for Bioethics for their support and guidance; the members of the Human Values project for thought-provoking discussions; and all the reviewers who contributed to this study, especially Cynthia Geppert, Elliott J. Crigger, Nada Salem, and Ihuoma Njoku.

## Supplementary Material

### Appendix A. Supplementary figures

Figures A1 to A6 support sections 8.6, 8.10 and 8.12. Panels A1 to A3 are regenerated on the harmonized scale described in section 8.1, and their score positions correspond to the values reported in the main text.

#### A.1 Distributional divergence

**Figure A1.**
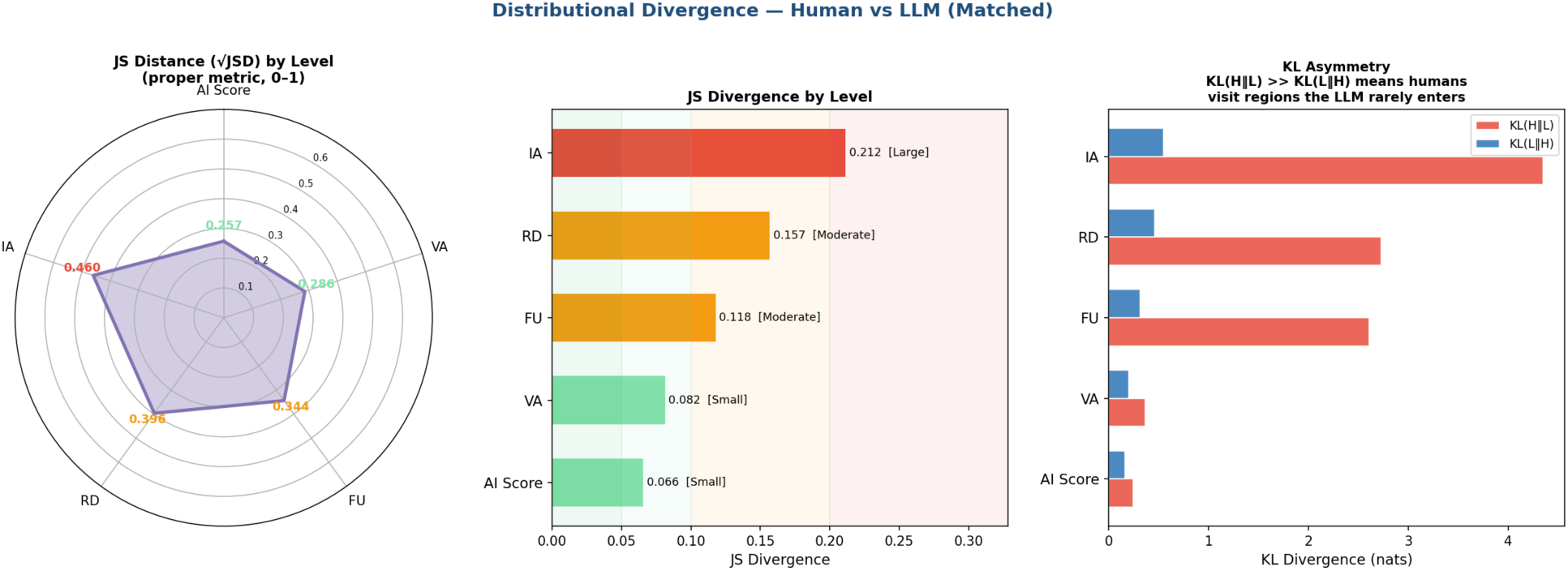
Distributional divergence between human and model score distributions at composite and domain level. Left: Jensen–Shannon distance, the square root of the divergence and a proper metric on the unit interval. Centre: Jensen–Shannon divergence with severity bands. Right: the two directed Kullback–Leibler divergences. The asymmetry is the informative panel: within the three domains carrying the largest mean differences, KL(human ǁ model) exceeds KL(model ǁ human) by roughly an order of magnitude, which indicates that human ratings occupy score regions the models enter rarely rather than the reverse. At the composite level the two directions are close to symmetric.

**Figure A2.**
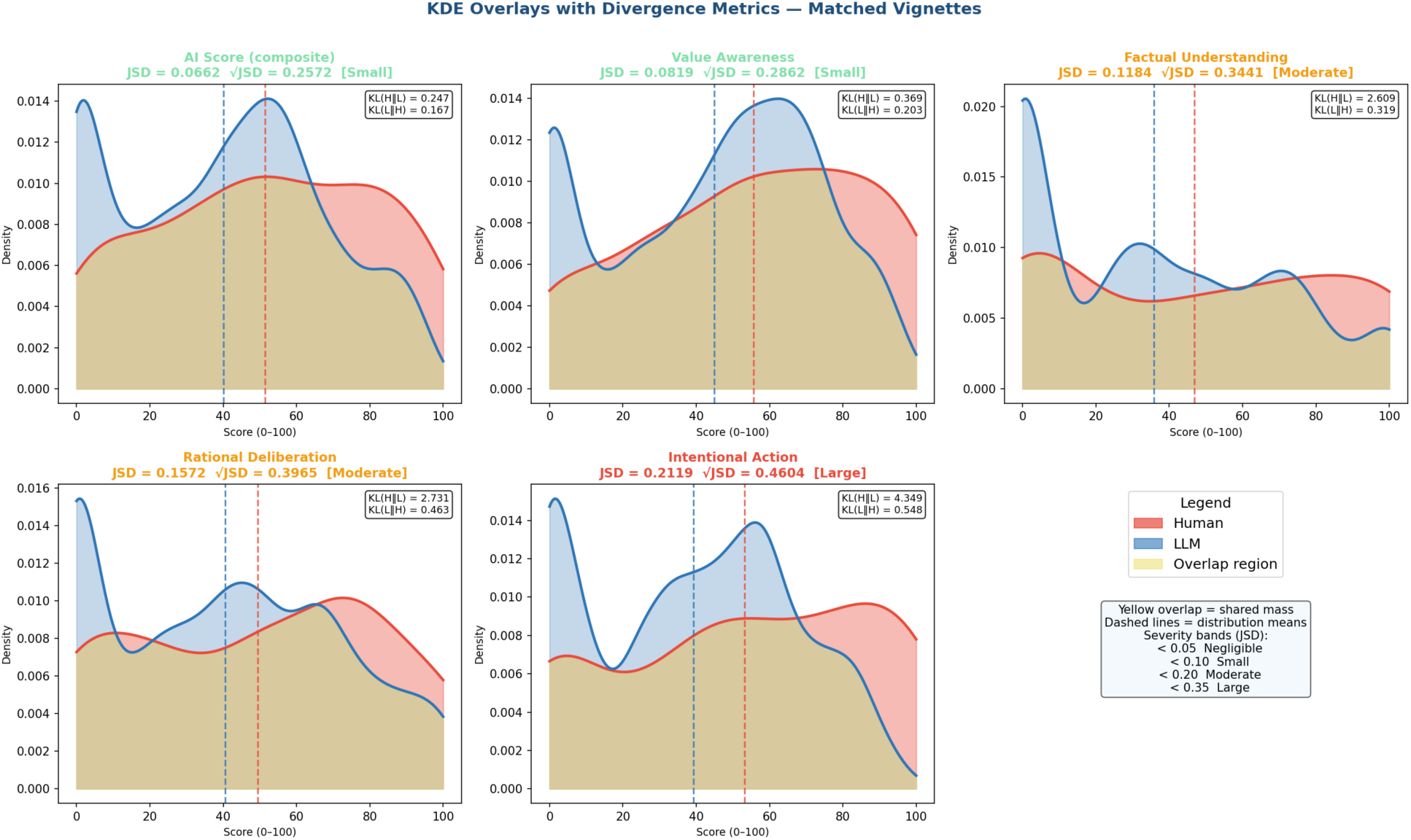
Kernel density overlays of human and model score distributions for the composite and each of the four domains, with the shared mass shaded. Dashed vertical lines mark distribution means. The model distributions are bimodal at every level, with one mode near zero and a second in the middle of the range, while the human distributions are broader and less clearly bimodal. The bimodality is the feature a mean difference conceals: models are not uniformly conservative but split between cases they score near the floor and cases they score moderately.

**Figure A3.**
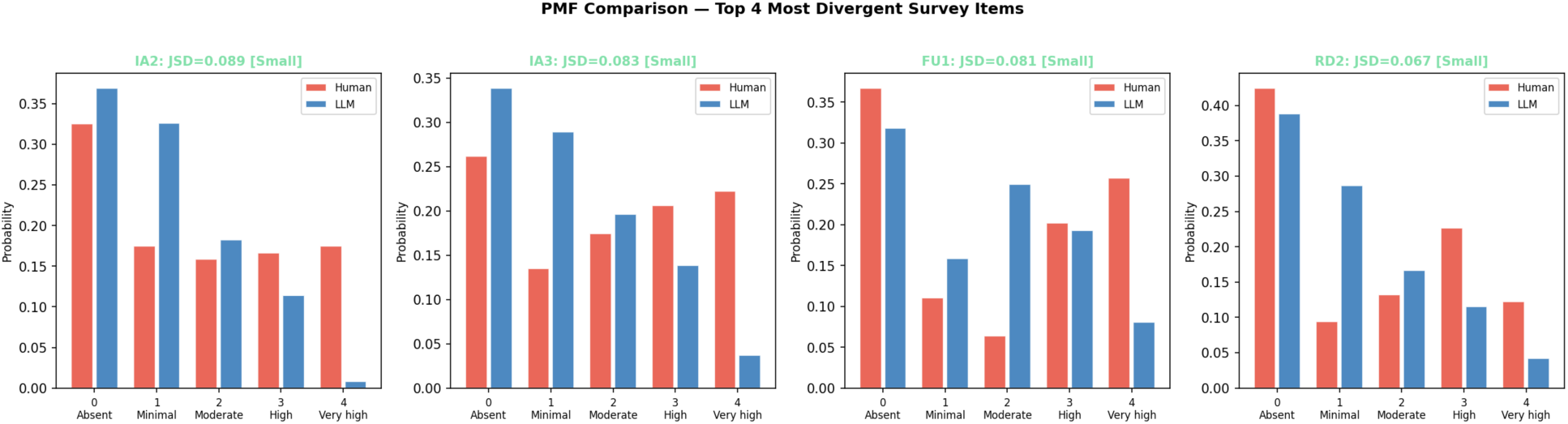
Response probability mass functions for the four instrument items with the largest Jensen–Shannon divergence. The distributions differ through modest shifts across several response categories rather than through a single uniform pattern. Human reviewers place more mass than models in the very high category for all four items, while both sources place substantial mass in the absent category. Differences also occur in the minimal and moderate categories. All four item-level divergences remain within the small band.

#### A.2 Per-vignette comparison

The following two figures report the human–model difference for each vignette individually, restricted to the vignettes with more than one human rating. They are included for completeness: the aggregate statistics in section 8 describe central tendency, and a reader assessing whether the difference is driven by a subset of cases needs the per-case view to check.

**Figure A4.**
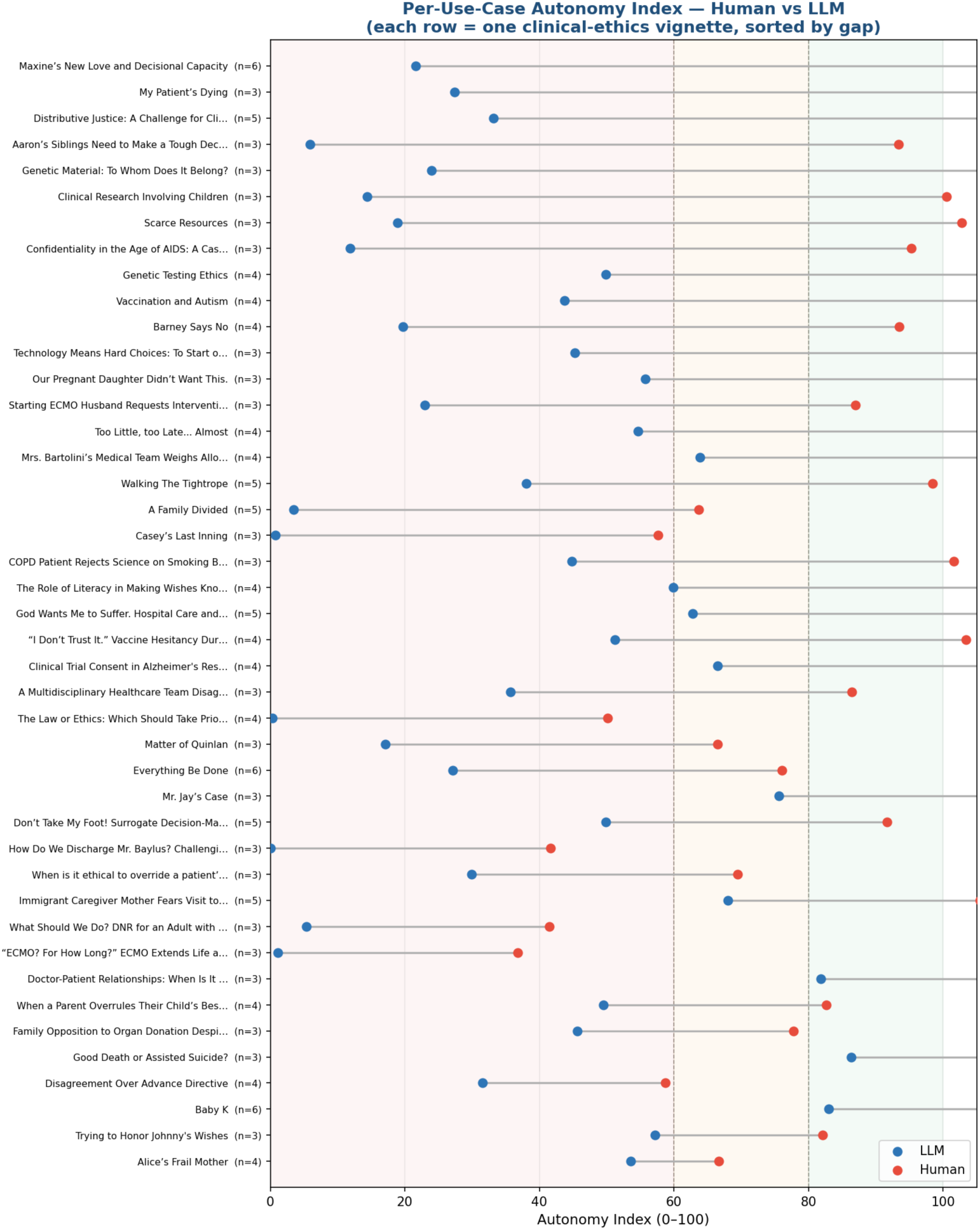
Human and model composite scores for each vignette, sorted by the size of the difference, with the number of human ratings in parentheses. Shaded bands mark the three interpretive ranges. The difference is present across the full corpus rather than concentrated in particular cases, which is consistent with the small rater-by-vignette interaction term reported in section 8.8.

**Figure A5.**
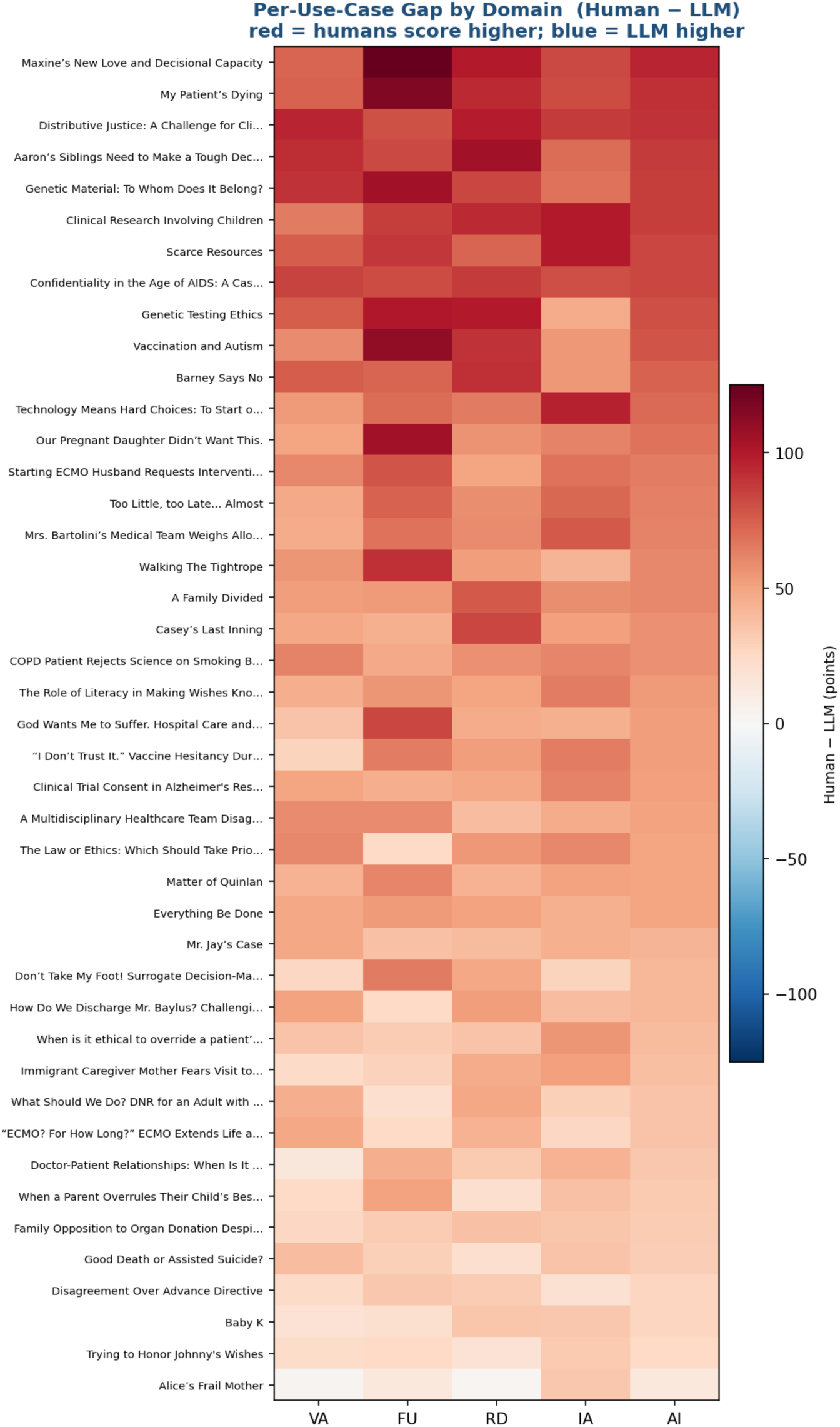
Human minus model difference by vignette and domain. Rows are ordered by mean difference. Fourteen of the fifty vignettes are negative at the composite level, and the domain pattern within a vignette is generally uniform, indicating that where models diverge from human reviewers they tend to do so across the construct rather than on one component.

**Figure A6.**
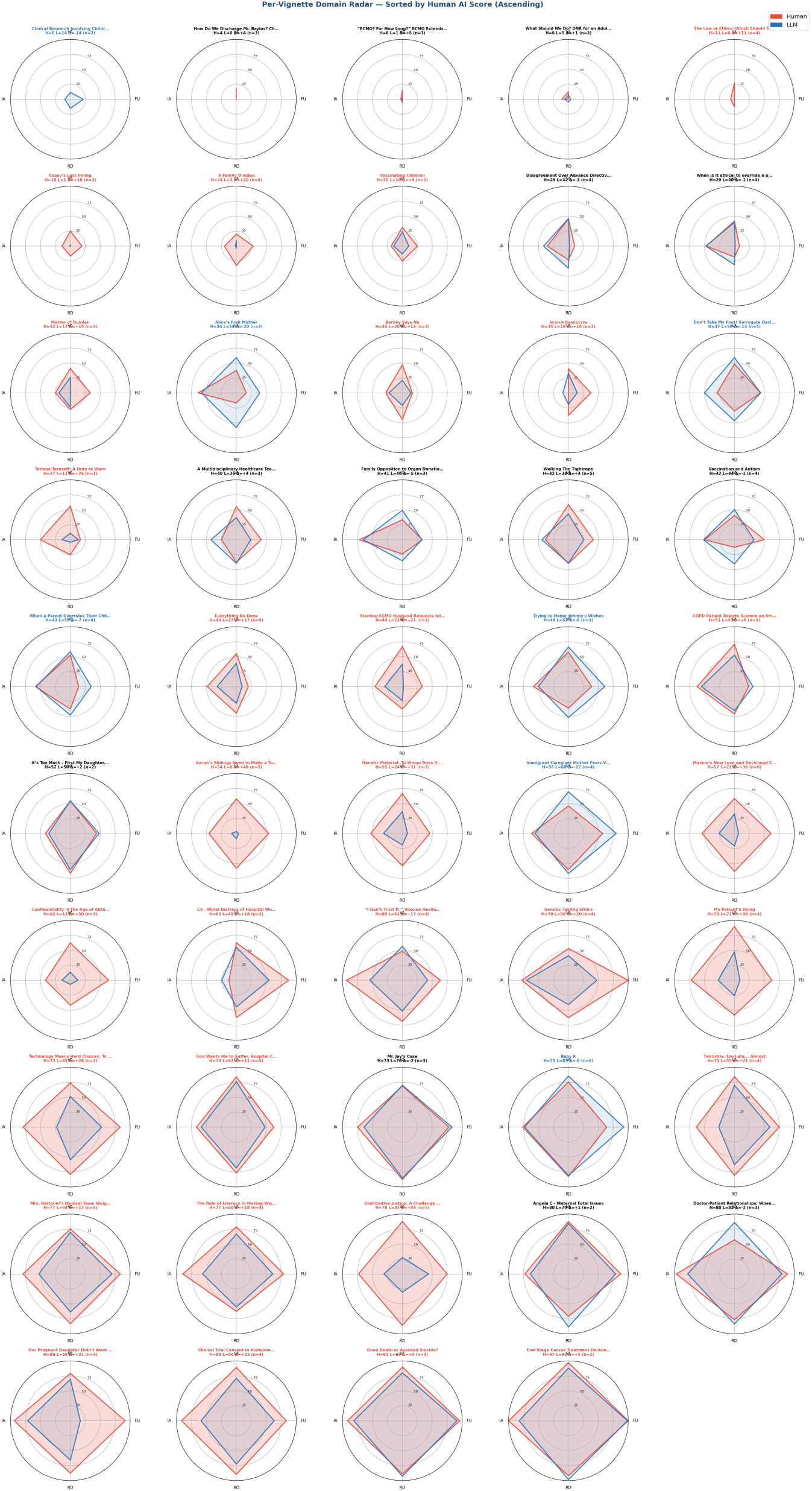
Radar chart per vignette comparing human and LLM responses.

